# Exclusive Enteral Nutrition Initiates Individual Protective Microbiome Changes to Induce Remission in Pediatric Crohn’s Disease

**DOI:** 10.1101/2023.12.21.23300351

**Authors:** Deborah Häcker, Kolja Siebert, Byron J Smith, Nikolai Köhler, Alessandra Riva, Aritra Mahapatra, Helena Heimes, Jiatong Nie, Amira Metwaly, Hannes Hölz, Quirin Manz, Federica De Zen, Jeannine Heetmeyer, Katharina Socas, Giang Le Thi, Chen Meng, Karin Kleigrewe, Josch K Pauling, Klaus Neuhaus, Markus List, Katherine S Pollard, Tobias Schwerd, Dirk Haller

## Abstract

Exclusive enteral nutrition (EEN) is the first-line therapy for pediatric Crohn’s disease (CD), but protective mechanisms remain unknown. We established a prospective pediatric cohort to characterize the function of fecal microbiota and metabolite changes of treatment-naïve CD patients in response to EEN. Integrated multi-omics analysis identified network clusters from individually variable microbiome profiles, with *Lachnospiraceae* and medium chain fatty acids as protective features. Bioorthogonal non-canonical amino acid tagging selectively identified bacterial species in response to medium chain fatty acids. Metagenomic analysis identified high strain-level dynamics in response to EEN. Functional changes of diet-exposed fecal microbiota were further validated in a combined approach using gut chemostat cultures and microbiota transfer into germ-free *Il10*-deficient mice. Dietary model conditions induced individual patient-specific strain signatures to prevent or cause IBD-like inflammation in gnotobiotic mice. Hence, we provide evidence that EEN therapy operates through explicit functional changes of temporally and individually variable microbiome profiles.

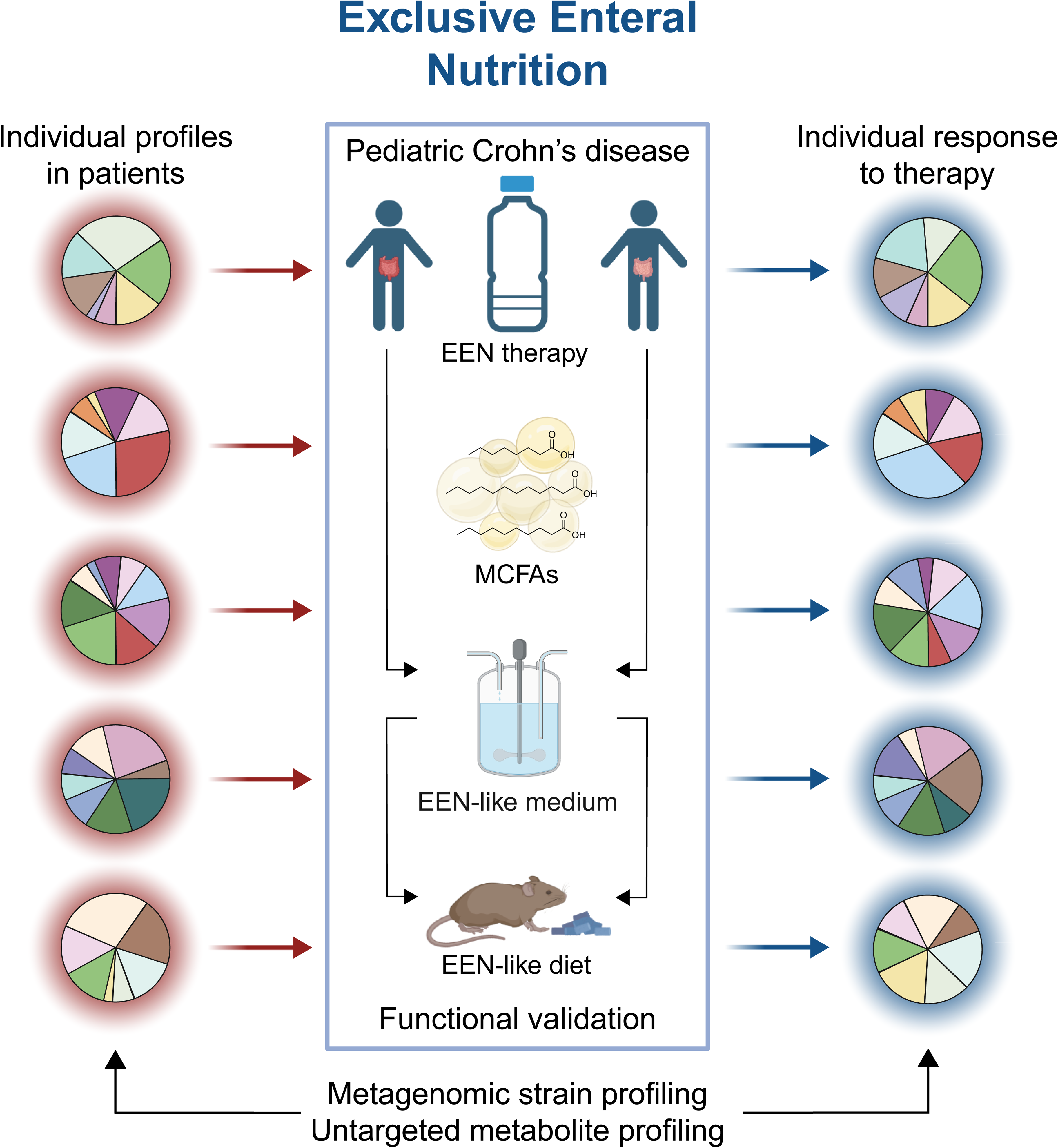

## Introduction

Crohn’s disease (CD), one of the main entities of inflammatory bowel diseases (IBD), is a chronic and relapsing inflammatory condition of the gastrointestinal tract^1^. Epidemiological studies have demonstrated a significant increase in the incidence of CD in Westernized countries, particularly among the pediatric age group^2,3^. The pathogenesis of IBD involves host genetics and environmental influences, among which diet and the intestinal microbiome are key etiologic factors^4,5^.

Exclusive enteral nutrition (EEN) is the first-line treatment for pediatric CD^6,7^ and also shows efficacy in adult patients^8^. EEN refers to polymeric liquid formulas, which are mostly devoid of fibers^9^. Re-introducing regular diet after 6-8 weeks of EEN often results in recurrence of inflammation and disease activity^10^. EEN has been shown to drive changes in gut microbiota composition^11^, but the high relapse rate after cessation of EEN suggests the presence of a CD-conditioned microbiome, which is able to re-establish after dietary intervention^12^. CD is associated with reduced bacterial diversity, compositional and functional changes, and the disruption of metabolic circuits between microbiome and host^13–15^. Despite recent findings that EEN mediates protective functions in experimental models of colitis^16^, human studies revealed incomplete recovery of the intestinal microbiome after antibiotic interventions^17^. Thus, functional evidence for EEN-mediated changes in the human microbiome as an underlying cause of therapeutic efficacy is still lacking.

Here, we identify EEN-protective signatures using integrated microbiota and metabolite analysis of treatment-naïve pediatric CD patients. Medium chain fatty acids (MCFAs) directly target bacteria from of the EEN-protective signatures, providing a direct mechanistic link between diet and microbiota changes. Further, we established a combined gut chemostat-to-mouse transfer approach to functionally validate EEN-mediated protective changes in the microbiome of CD patients. We demonstrate that EEN directly shapes microbiomes to mediate protective functions in treatment-naïve CD patients, operating through explicit patient-to-patient changes of temporally and individually variable strain profiles.

## Results

### Microbiota and metabolite profiling identify individual responses to EEN therapy in pediatric CD patients

We established a pediatric IBD cohort and longitudinally followed 78 IBD patients, showing no disease specific but individual bacterial and metabolite profiles in fecal samples (**Fig. 1A, S1A-C**). To gain insides in mechanisms of EEN therapy, we focused explicitly on 20 newly diagnosed and treatment-naïve CD patients, receiving EEN as induction therapy (CD-EEN group, 9 females, age at study inclusion 12.5 ± 3.1 years) (**Fig. 1A, S1A; Table S1**). EEN was highly effective and significantly improved disease activity and inflammatory markers at the end of EEN treatment, resulting in clinical response and induction of remission in 20/20 and 18/20 patients, respectively (**Fig. 1B**). During follow-up, patients received methotrexate (6/20), anti-TNF (13/20), or combination therapies (9/13) as maintenance treatment. Relapse after EEN was observed in 8/20 patients over one-year follow-up (**Fig. 1B**). Two patients switched from Modulen IBD® to Neocate Junior® during EEN due to suspected cow’s milk protein allergy.

**Figure 1:**
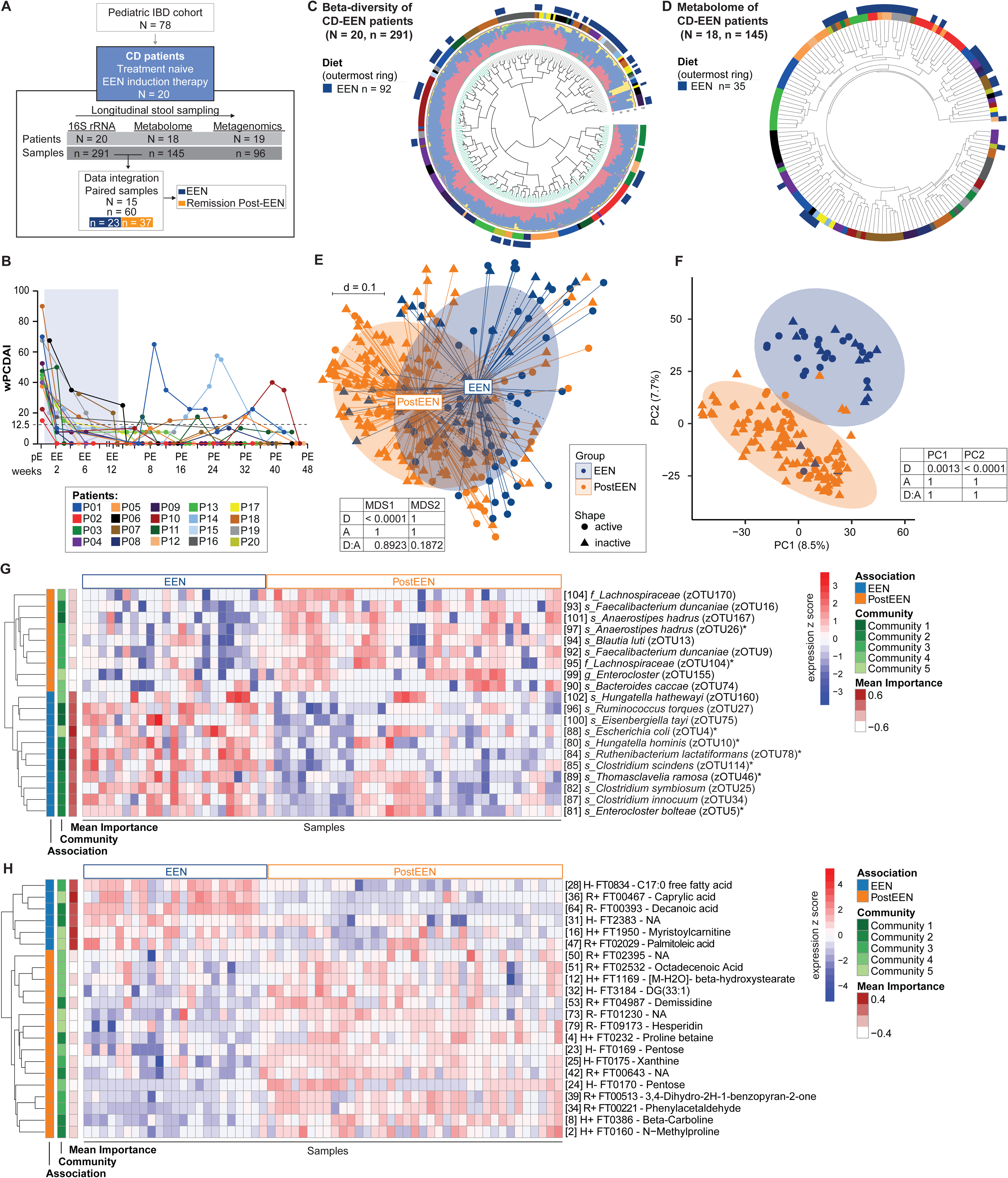
Fecal microbial and metabolic changes in response to EEN. Patients of the Pediatric IBD cohort with focus on 20 newly diagnosed and treatment naïve CD patients, receiving EEN induction therapy. Longitudinal collected samples (n = samples, N = patients) that were used for 16S rRNA amplicon and metagenomic sequencing as well as metabolome analysis are shown. Paired samples (n = 60) were used for data integration of 16S rRNA and metabolome data (samples highlighted in blue = taken under EEN, orange = taken PostEEN). **B** Weighted pediatric Crohn’s Disease activity index (wPCDAI) over time (EEN shaded blue), remission achieved below 12.5 points (dashed line). pE = PreEEN, EE = EEN, PE = PostEEN. **C** Beta-diversity analysis of the fecal microbiota samples. The circular dendrogram shows differences in beta-diversities from the longitudinal microbial profiling based on generalized UniFrac distances between the 291 stool samples of the 20 patients. Indicated in color code are the patients and samples collected during EEN. Color code of taxonomic composition at phylum level as in B. Branch color indicates results of unsupervised clustering. Patient colour code found in B. **D** Visualization of untargeted metabolome samples from 18 newly diagnosed pediatric CD patients receiving EEN therapy: Complete clustering with Euclidean distance of longitudinal samples from 18 CD-EEN patients (n = 145), Patient colour code found in B. **E** Beta-diversity MDS plot samples during EEN (n = 89, active: 55, inactive: 34) vs. PostEEN (n = 188, active: 41, inactive: 147). **F** Analyses of untargeted metabolomics with representative PCA plot comparing samples during EEN (n = 35, active: 21, inactive: 14) and PostEEN (n = 106 active: 21, inactive: 85). Axes indicate principal components (PC), with PC1 representing the most variation (%) and PC2 representing the second most variation (%). (**E, F**) Coefficients were tested with a t-test, reported p-values are Bonferroni-corrected, D = diet, A = activity, D:A = interaction term between diet and activity. **G, H** Heatmaps depicting the abundances of the top selected feature from the correlation network Supplementary Fig. S2. Selected sPLS-DA features were subsetted by their mean importance (see Star Methods: Data integration of 16s rRNA amplicon data with metabolomics and metagenomics for details). All values shown are feature-wise z-scores. Information on association with EEN (blue), PostEEN (orange), sample collection under which diet (EEN blue), PostEEN (orange), community association (1 – 5) are indicated; **G** zOTUs,*BONCAT responders Fig.2, **H** Metabolites.

Analysis of samples before (PreEEN), during (EEN), and after EEN therapy (PostEEN) (microbiota profiling: 291 samples, metabolomic analysis: 145 samples) revealed patient-specific responses (**Fig. 1C, D**). Bacterial richness based on zOTUs (zero-radius operational taxonomic units) changed individually during formula feeding (**Fig. S1D**) but was lower during active CD compared to remission (**Fig. S1E**). Samples of EEN-treated patients are scattered throughout dendrograms of microbiota and metabolome profiles (**Fig. 1C, D**). Despite considerable overlap in multi-dimensional scaling (MDS) plots based on microbial profiles, linear mixed models (LMMs) identified only diet as significantly contributing to the first MDS dimension (x-axis) (**Fig. 1E**, n=277, p-value < 0.0001; **Fig. S1F**). PCA plots of metabolic profiles also showed significant separation according to diet along PC2 but no separation according to disease activity (**Fig. 1F**, n = 141, p-value < 0.0001; **Fig. S1G**).

### Integration of microbiota and metabolome data identifies key features of EEN exposure

To identify key differences between fecal microbiota and metabolite profiles of patients collected under EEN (therapy, partially with still active disease) and PostEEN conditions (receiving regular diets but completely in remission), a network analysis incorporating matched zOTU taxonomic profiles and untargeted metabolomics from 60 samples was performed. The identified interlinked features clustered in five distinct network communities (**Fig. S2**). In summary for the top selected features from the correlation network, higher mean importance was found for zOTUs associated to EEN compared to zOTUs associated with PostEEN (**Fig. 1I**). The top eleven zOTUs associated to EEN were found within all communities (**Fig. 1I**, green gradient), including several zOTUs from the genera *Enterocloster* and *Clostridium* (formerly *Lachnoclostridium; Lachnospiraceae;* **Table S2**) which still showed individual abundances in each patient (**Fig. S3A**). Of the 22 top selected metabolite features, the majority were associated with PostEEN, whereas six metabolites were associated with EEN with higher mean importance (**Fig. 1J**). Of the in total ten metabolites associated with EEN, the majority of those were medium chain fatty acids (MCFA) (palmitoleic acid, decanoic acid, caprylic acid and lauric acid) which were identified as metabolite features present in Modulen IBD® (**Fig. S2C, S3B**).

### Medium chain fatty acids selectively induce translational activation of fecal bacteria

Three MCFAs [(lauric acid (LA), decanoic (DA) acid and octanoic acid (OA)] were identified as signature metabolites of EEN, likely originating from IBD Modulen. To determine which gut bacteria can be activated by these metabolites, we employed a combination of bioorthogonal noncanonical amino acid tagging (BONCAT) and fluorescence-activated cell sorting (FACS) to identify translationally active bacteria in the presence of MCFAs under *ex vivo* anaerobic incubations (**Fig. 2A**). We selected three donors (MP1-MP3, model patient) within the CD-EEN group based on clinical activity, limited drug exposure and chose four different samples along EEN therapy (**Table S3,** STAR Methods, **Fig. 4A, C, I; S8F**). Confocal microscopy visualization and quantification indicated that a significantly higher proportion of gut bacteria became translationally active after exposure to MCFAs, compared to the solvent control (**Fig. 2B, C; Fig. S4, S5**). The MCFA-active fraction of the community (BONCAT labeling and FACS sorting), exhibited significant differences compared to the unsorted community and between patients as shown in the MDS ordination (p = 0.001, **Fig. 2D**). Ethanol mediated variability is shown as additional control (**Fig. S5C**). This separation highlights distinct microbial communities to be present after MCFAs exposure. We identified different zOTUs enriched in the active fraction after exposure to MCFAs compared to the ethanol control (**Fig. 2E, S5; Table S4, S5**). Different species from the *Enterocloster* genera were significantly enriched in response to MCFAs, especially in MP2 at EEN remission stage (**Fig. 2E**). This includes *Enterocloster aldensis* and two subspecies of *Enterocloster boltae*, which aligns with the bacterial signature identified from the clinical cohort (**Fig. 1G**). Additional enriched taxa included *Eggerthella lenta* in MP2 (remission); *Escherichia coli, Clostridium scindens, Dialister invisus, Sporobacter* spp. *and Eubacterium* spp. (MP2 relapse); *E. coli* in MP1 and MP3 as well as *Lachnospiraceae* in MP1. In contrast, several additional zOTUs were depleted in the active fraction after MCFA exposure including *Clostridium scindens, Thomasclavelia ramosa, Hungatella hominis, Peptostreptococcus anaerobius, Ruthenibacterium lactatiformans* and *Phocaeicola vulgatus* in MP2 at EEN remission stage and *Anaerostipes hadrus* and *Hominenteromicrobium mulieris* in MP2 under relapse (**Fig. 2E, S5, Table S4, S5**). In summary, of the 16 zOTUs associated with EEN from the Network analyses, 8 zOTUs responded to MCFA exposure (labeled with * in **Fig. 1G, S2C**).

**Figure 2:**
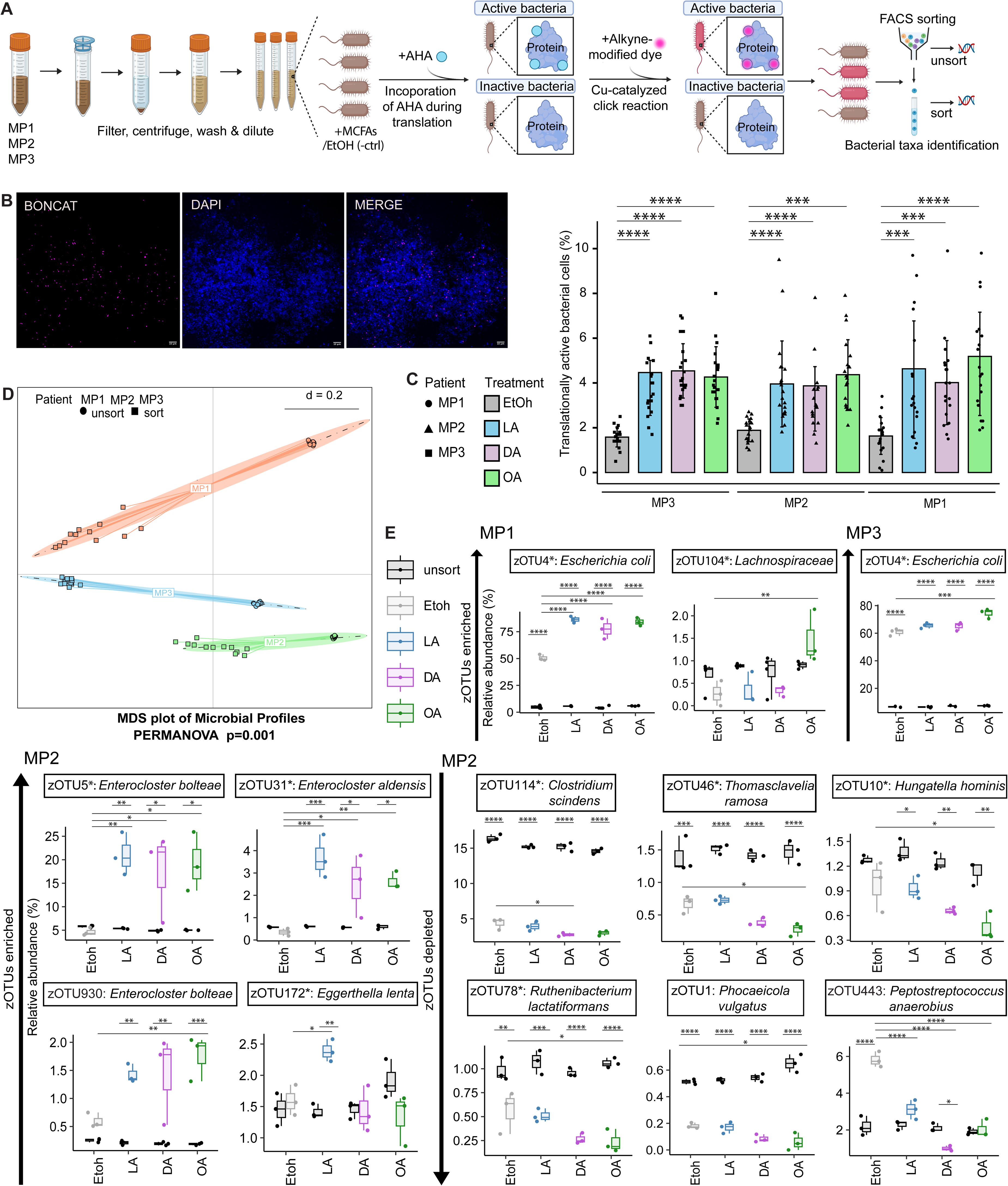
MCFAs-Induced Activation of Translationally Active Gut Bacteria. **A** Experimental design. MP1, MP2 and MP3 stool samples previously stored in glycerol, were incubated under anaerobic conditions and in the presence of medium chain fatty acids (MCFA)[(lauric acid (LA), decanoic acid (DA) and octanoic acid (OA)] and of the cellular activity marker (L-Azidohomoalanine (AHA). Translationally active bacterial cells labelled with alkyne-modified dye were sorted with BONCAT-FACS. The taxonomic profiles of sorted and unsorted cells were identified by 16S rRNA genes amplicon sequencing. **B** Representative confocal microscopic images of one stool samples (MP2) stimulated with OA. Pink: active cells (BONCAT-Cy5); Blue: all cells (DAPI). Scale bar is 10µm. **C** Translationally active bacterial cells quantified in MP1, MP2 and MP3 patients. P values were calculated by Kruskal-Wallis test and Dunn test for multiple comparisons. *p < 0.05, **p < 0.01, ***p < 0.001, ****p < 0.0001. **D** Beta-diversity MDS plots shows clustering of samples based on model patients and sorted-unsorted groups (perMANOVA p=0.001 for both comparisons). MP1 = orange, MP2 = green, MP3 = turquoise, sorted cells = square, unsorted cells = circle. **E** Relative abundances of significant enriched and depleted zOTUs in MP1, MP2 and MP3. *zOUTs identified in Network analyses Fig 1G, S2. P values were calculated by ANOVA and Tukey’s test for multiple comparisons. *p < 0.05, **p < 0.01, ***p < 0.001, ****p < 0.0001.

### Microbial species, strains, and functions change dramatically at EEN cessation

Considering the various changes of community profiles associated with EEN or PostEEN, we next investigated dynamics within the patients along EEN therapy. Cessation of EEN resulted in an elevated turnover in zOTU composition (**Fig. 3A**), resulting in higher dissimilarity - even after controlling for greater time periods - between pairs of samples spanning the transition off of EEN compared to during or post-EEN alone (p < 10^−3^ for both comparisons by permutation test). Using shotgun metagenomes for 96 CD-EEN patient samples (n = 19 patients), we applied StrainFacts^18^ to resolve taxonomic differences below the level of species. Like zOTUs, and despite the much smaller number of comparisons, we detected the same elevated turnover in the composition of strains during the transition off of EEN relative to the PostEEN period (p < 10^−3^, **Fig. 3B**). Overall, we observed high strain-level diversity within and between CD-EEN patients. For most of the 369 species, individual strains were largely specific to a single patient and appeared in multiple samples from their host, while some samples contained multiple distinct strains of a given species (e.g. **Fig. 3D-F**).

**Figure 3:**
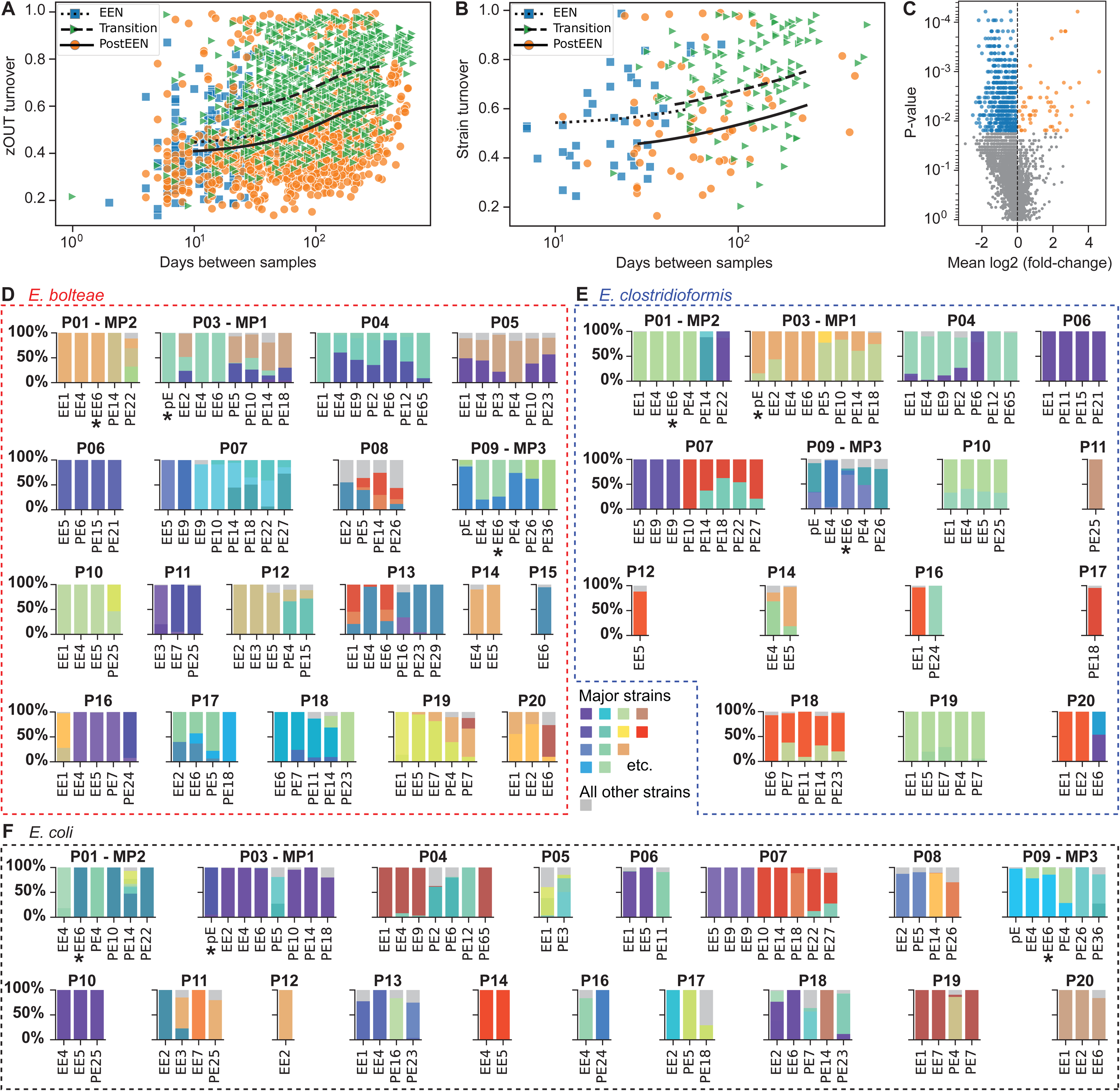
Species and strain turnover during the transition off of EEN: **A,B** Turnover of taxonomic composition during the transition from EEN to a PostEEN diet of both **(A)** zOTU and **(B)** strain compositions. The three lines reflect cubic spline regression of Bray-Curtis dissimilarities against time, averaged across subjects and offset by an independent intercept for each pair type. **C** Volcano plots showing COGs with significant changes in normalized abundance at the transition off of EEN (Wilcoxon test at false discovery rate (FDR) < 0.1). Here, 54 COGs were at significantly greater relative depth during PostEEN (orange points) and 529 during EEN (blue). **D-F**. Composition over time of strains within **(D)** E. bolteae (red box), **(E)** E. clostridioformis (blue box), **(F)** E. coli (black box) all three abundant in the CD-EEN patients: All three species have notable strain dynamics during and after EEN. Strain genotypes and fractions were estimated from shotgun metagenomic data. Fractions of abundant strains (colored bars) are shown across samples from individual subjects (P01, P02…), before EEN (“pE” labels on x-axis), during (“EE”) and after (“PE”). *Samples used as donor samples in other experiments. Strain fractions for the species sum to 1; minor strains are summed together and shown in grey. An analogous plot for B. uniformis is in Supplementary Figure S4D.

Besides taxonomic turnover, we also detected 583 protein coding gene families (COGs) with substantial changes in their normalized abundance (54 increased and 529 decreased) at the transition off of EEN (**Fig. 3C**). Families increased during EEN included COG3845 (mean log2 PostEEN/EEN of −0.58) and COG4603 (−0.58), components of the ABC-type guanosine uptake system. Increases in COG4813 (+2.1), annotated as trehalose utilization protein, and COG3867 (+1.0), arabinogalactan endo-1,4-beta-galactosidase, indicate that microbes encoding these proteins respond to host diet and may play a role in metabolite transformations. These findings are consistent with changes in carbohydrates and sugar acids found in the metabolite analysis (**Fig. S2C**).

Next, we sought to identify species with high prevalence and abundance and with large changes in relative abundance or extensive strain turnover at the end of EEN. Using an indicator score based on these features (**Fig. S2A, Table S7**), we flagged 46 species as highly dynamic. We identified pairs of zOTUs and metagenomic species (**Table S6**) and matched 37 (80%) of the top scoring species (**Fig. S6B, Table S7**). Here we highlight and contrast the strain dynamics of two closely related species, *Enterocloster bolteae* (formerly *Lachnoclostridium bolteae*) and *E. clostridioformis* (formerly *C. clostridioforme*). Both of these species were matched to zOTU5 (**Fig. S6C**) and showed dynamic strain composition over patient time series (**Fig. 3D, E**). In some patients, EEN and PostEEN samples contained similar strains of each species, but the dominant strain changed; in others, a new strain appeared PostEEN that was absent or undetectable during EEN. Interestingly, the relative abundance of *E. bolteae*, but not *E. clostridioformis*, was significantly affected by EEN cessation (p = 0.018 and 0.9, respectively, by Wilcoxin test; **Table S7**). Hower, while both showed a similar strain turnover overall, *E. clostridioformis*, but not *E. bolteae* had significantly elevated strain turnover at the transition from EEN to PostEEN (p = 0.004 and 0.202, respective, by permutation test: n=999). This effect was also seen in nine other common species (**Table S7**), including *E. coli* (**Fig. 3F**). By contrast, some species like *Bacteroides uniformis* revealed notably lower strain diversity within samples and across time (**Fig. S6D**).

The distinction in response to EEN among strains and between closely related species reinforces the importance of strain-level characterization in individuals. Altogether, metagenomic analyses demonstrate that gut microbial communities are dynamic at and below the species level and highly influenced by EEN, motivating us to explore their relationship to the clinical efficacy of EEN.

### EEN-like diet triggers protective changes in fecal microbiota

To further evaluate the functional impact of EEN on patient-derived fecal microbiota, we used continuous culture systems in a gut chemostat model and fecal microbiota transfer (FMT) into germ-free (GF) WT and *Il10*^−/−^ mice. *Il10*^−/−^ mice are a microbiota-dependent IBD mouse model responsive to human fecal microbiota, as previously shown for adult CD patients^19^ (**Fig. 4A**). The previously used samples from model patients (MP) for the BONCAT experiments were also selected for the transfer experiment (**Fig. 4B, C, I**; **Table S3**, STARMethods).

**Figure 4:**
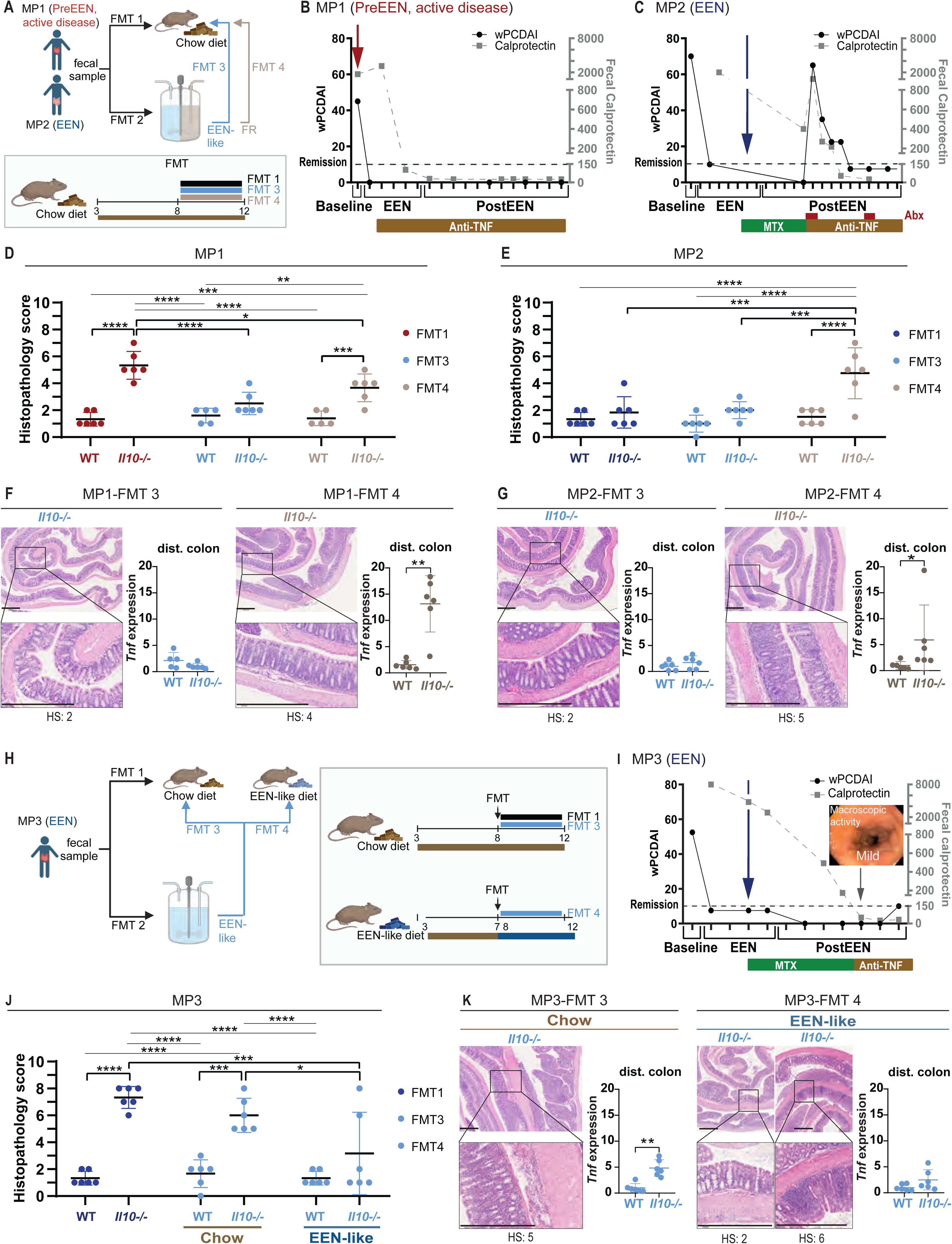
Functional validation of gut microbiota-dependent EEN-efficacy using *ex vivo* fermentation and gnotobiotic humanized mice. **A** Experimental scheme: FMT 1, Patient sample is transferred into GF mice at the age of 8 weeks for 4 weeks. Same patient sample is also transferred (FMT 2) in an **ex vivo** gut chemostat model and treated with medium simulating EEN (EEN-like, devoid of fibers) or a regular diet (fiber-rich, FR). Fermented sample is transferred (post *ex vivo*) into GF mice (age of 8 weeks) for 4 weeks (FMT 3 - EEN-like medium conditioned, FMT 4 - FR medium conditioned). **B, C** Transfer of patient sample: patients disease state with wPCDAI and fecal calprotectin levels, therapy response over time with maintenance therapy (MTX, Anti-TNF) and sampling time point. **B** Model Patient (MP) 1, sample (red arrow) collected PreEEN in active disease. **C** MP2, sample (blue arrow) collected in EEN induced remission. **D** Mouse histopathology scoring of colon Swiss roles of FMT 1, 3 and 4 from MP1. **E** Mouse histopathology scoring of colon Swiss roles of FMT 1, 3 and 4 from MP2. **F, G** Mouse transfer experiments from FMT 3 and 4: Representative (mean of group) H&E stained sections of colonic Swiss roles and corresponding higher magnifications for Il10^−/−^ mice (scale bars = 500 μM, HS = histopathology score) respective controls in Extended Data Fig 3) with *Tnf* gene expression levels (fold of control) in distal colon. **F** MP1. **G** MP2. **H** Experimental scheme: Patient sample from MP3 is transferred into GF mice (FMT 1) at the age of 8 weeks, for 4 weeks. Same patient sample is also transferred in an *ex vivo* gut chemostat model and treated with EEN-like medium. Fermented sample is transplanted (post *ex vivo*) into GF mice at the age of 8 weeks, for 4 weeks getting either chow diet (FMT 3) or EEN-like purified diet (EEN-like) one week prior to FMT (FMT 4). **I** MP3 sample for transfer (EEN): patients’ disease state with wPCDAI and fecal calprotectin, therapy response over time, maintenance therapy (MTX, Anti-TNF) and sampling time point of transferred fecal sample (blue arrow); grey arrow indicates endoscopy performed in the patient PostEEN and the picture taken during endoscopy shows active ileitis. **J** Mouse histopathology scoring of colon Swiss roles of FMT 1, 3 and 4 from MP3. **K** Mouse transfer experiments from FMT 3 and 4 from MP3: Representative (mean of group) H&E stained sections of colonic Swiss roles and corresponding higher magnifications for Il10^−/−^ mice (scale bars = 500 μM, HS = histopathology score) respective controls in Extended Data Fig 3) with *Tnf* gene expression levels (fold of control) in distal colon. **A, H** Created with BioRender.com. Data are represented by (**D, E, F, G, J, K**) mean ± SD of six biological replicates. P values were calculated by Mann-Whitney test (**F, G, K**) or two-way ANOVA with Tukey multiple pairwise-comparisons test (**D, E, J**). *p < 0.05, **p < 0.01, ***p < 0.001, ****p < 0.0001

Microbiota transfer of the PreEEN sample of MP1 (MP1-FMT 1, **Fig. 4B**, red arrow) induced colonic tissue inflammation in all gnotobiotic *Il10*^−/−^ mice (histopathology score of 5.3 ± 1.0) but not in WT control mice (**Fig. 4D** p < 0.001, histopathology score of 1.3 ± 0.5; **Fig. S7A** H&E), mirroring the active disease state of this patient. Following transfer of MP1 fecal material into the gut chemostat model (MP1-FMT 2, **Fig 4A**), dietary exposure was mimicked using combinations of fiber-free EEN-like (including 10% Modulen IBD® of the 1 kcal/mL preparation) and fiber-repleted (FR) medium, respectively (**Fig. S8A, B**). We then transferred microbiota (MP1-FMT 3) from the chemostat model exposed to EEN-like medium and thereby reversed the inflammatory phenotype of MP1 in *Il10*^−/−^ mice (**Fig. 4D**, histopathology score of *Il10*^−/−^ mice = 2.5 ± 0.8; WT mice = 1.6 ± 0.5; FMT1 vs FMT 3 *Il10*^−/−^ p <0.001; **Fig. 4F** H&E, **Fig. S7A**). Conversely, under fiber-repleted medium conditions MP1-FMT 4 induced IBD-like tissue pathology in four out of six *Il10*^−/−^ mice (**Fig. 4D** histopathology score = 3.7 ± 1.0; **Fig. 4F** H&E, **Fig. S7A**), compared to disease-free WT mice (**Fig. 4D** histopathology score = 1.4 ± 0.5, p = 0.0087; **Fig. 4F** H&E, **Fig. S7A**). *Tnf* gene expression confirmed active tissue inflammation with significantly elevated expression levels in *Il10*^−/−^ compared to WT mice (**Fig. 4F** p = 0.0022 dist. (distal) colon; **Fig. S7D** p = 0.0152 prox. (proximal) colon). Thus, *ex vivo* EEN-like intervention recapitulated therapeutic functional changes to the microbiota.

Transfer of the fecal microbiota from MP2 failed to induce inflammation in *Il10*^−/−^ mice (MP2-FMT 1, **Fig 4E** H&E, **Fig. S7B**), resembling the inactive disease state of the patient. Since the stool sample of MP2 was retrieved under EEN therapy, we started the gut chemostat on EEN-like medium (MP2-FMT 2; **Fig. S8C**). In contrast to MP2-FMT 3 (**Fig. 4E** histopathology score in in *Il10*^−/−^ = 2.0 ± 0.6; WT = 1.0 ± 0.6; **Fig. 4G** H&E**, Fig. S7B**), transfer of FR-conditioned microbiota (MP2-FMT 4) triggered IBD-like colitis, including histopathologic changes and elevated *Tnf* tissue expression (**Fig. 4E**, histopathology score in *Il10*^−/−^ = 4.8 ± 1.9 *vs*. WT = 1.5 ± 0.5, p (*Il10*^−/−^ FMT1 *vs*. FMT4) = 0.0004, p (*Il10*^−/−^ FMT3 *vs*. FMT4) = 0.0008 ; **Fig. 4G** H&E, *Tnf* gene expression p = 0.0152 dist. colon; **Fig. S7D** p = 0.0022 prox. colon). Thus, both transfer experiments (MP2-FMT 1 and MP2-FMT 3) demonstrate protective effects of EEN. Reintroduction of fiber (MP2-FMT 4) reverted this protective function, indicative for the risk of relapse in MP2 after EEN cessation (**Fig. 4C**).

Next, we introduced an EEN-like mouse diet one week prior to FMT (MP3-FMT 4) to evaluate extended dietary effects in gnotobiotic WT and *Il10*^−/−^ mice (**Fig. 4H**). MP3 had elevated fecal calprotectin levels of 2828 mg/L (**Fig. 4I**, blue arrow) and ongoing mucosal inflammation with endoscopic signs of disease activity (**Fig. 4I**, grey arrow), despite achieving clinical remission under EEN therapy. Accordingly, FMT 1 from MP3 induced severe inflammation in all recipient *Il10*^−/−^ mice (**Fig. 4J** histopathology score in *Il10*^−/−^ = 7.3 ± 0.8 *vs*. WT = 1.3 ± 0.5, p = 0.0022; **Fig. S7C** H&E). In contrast to MP1 and MP2 (**Fig. 4D, E**), EEN-like medium in the chemostat model did not change the inflammatory phenotype in IBD-susceptible mice (MP3-FMT 3, **Fig. 4J** histopathology score in *Il10*^−/−^ = 6.0 ± 1.3 *vs*. WT = 1.6 ± 1.0, p = 0.0006; **Fig. 4K** H&E, **Fig. S7C**; *Tnf* p = 0.0022 for dist. colon and prox. colon; **Fig. S7D**). However, introducing EEN-like diet decreased the level of disease severity after MP3-FMT 4 in most *Il10*^−/−^ mice (**Fig. 4J** histopathology score in EEN-like-fed *Il10*^−/−^ = 3.2 ± 3.1 *vs*. WT = 1.3 ± 0.5, p = n.s.; **Fig. 4K** H&E, *Tnf* p = n.s dist. colon; **Fig. S7C, D,** *Tnf* p = 0.0022 for prox. colon) and significantly reduced disease activity compared to the chow-fed group (**Fig. 3J** FMT 3 *vs*. FMT 4, histopathology score in EEN-like-fed *Il10*^−/−^ = 3.2 ± 3.1, p = 0.0271, *Il10*^−/−^ FMT 1 *vs*. FMT 4 p = 0.0005, **Fig. 4K** H&E, **Fig. S7C**). An additional sample of MP2 after clinical relapse was tested under conditions of the extended EEN-like feeding (**Fig. S8F**, orange arrow), and contrary to MP3, the MP2 relapse sample was resistant to EEN induced protection (**Fig. S8E-L**). These findings suggest that sustained EEN-like feeding increased the chance to prevent recurrence, but efficacy of repeated EEN decreases with a second course.

### Individual strain signatures in gnotobiotic *Il10*^−/−^ mice convey protective function of patient’s microbiota in response to diet

To investigate bacterial strain profiles responsible for the mouse disease phenotypes (**Fig. 4**), we performed metagenomic analyses of all 54 gnotobiotic *Il10*^−/−^ mice colonized with fecal communities of MP1-MP3 (**Fig. 5A-C**, triangles: red - active disease, green - inactive disease; **Fig. S9A**). Out of 100 strains detected in MP1, 64 were specific to mice with active disease, while only three were specific to inactive disease (**Fig. 5A**, lowest Venn diagram (Vd)). Strains in MP2 mice were skewed towards being specific to inactive disease or shared (**Fig. 5B**, lowest Vd), consistent with lower disease activity in MP2 and fewer mice with active disease. Strains in MP3 mice were skewed towards being specific to active disease, probably due to a higher number of MP3 mice having active disease (**Fig. 5C**, lowest Vd).

**Figure 5:**
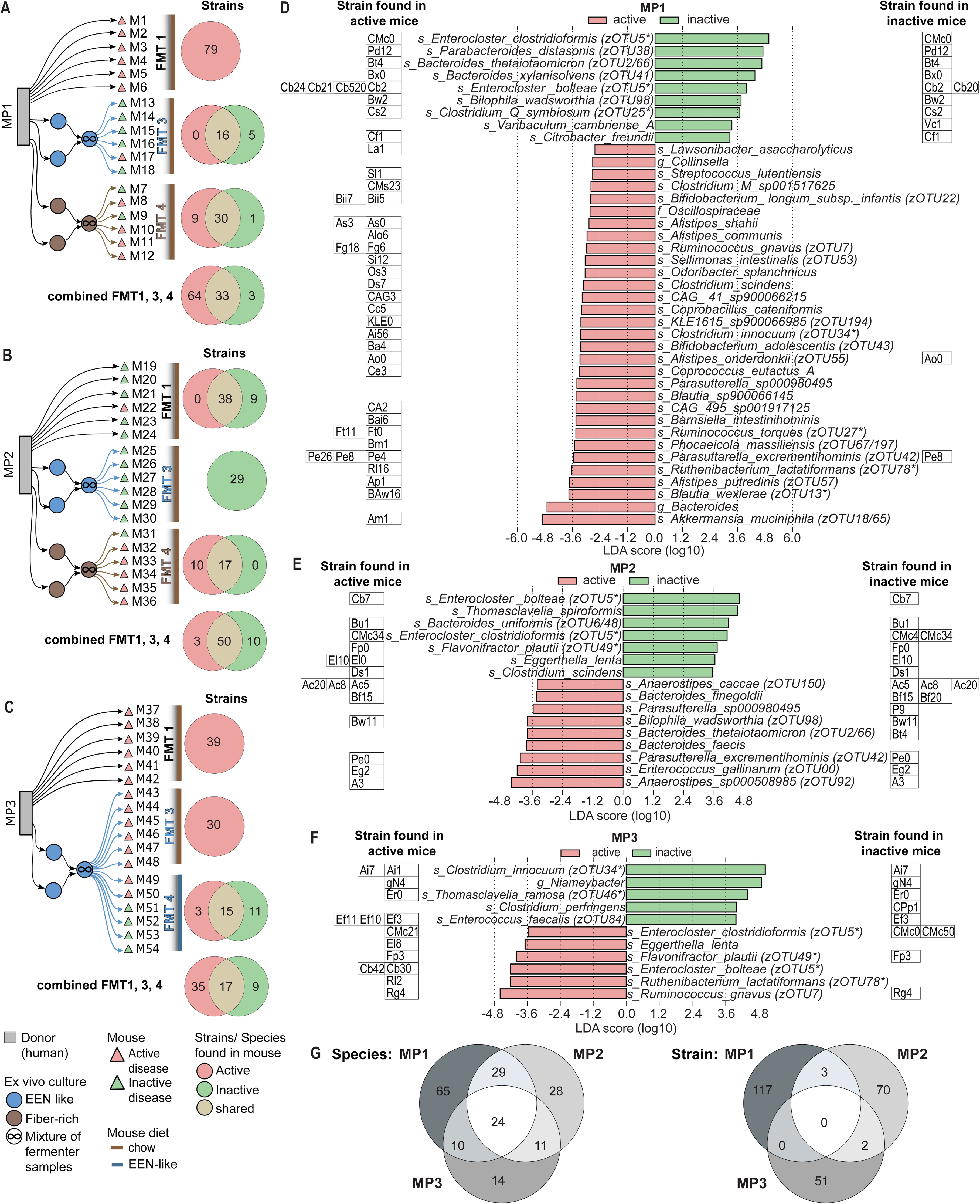
Strains associated with active or inactive disease in Il10^−/−^ mice: **A-C** Transfer overview of donor sample (grey) from FMT 1 (black), FMT 3 (EEN-conditioned, blue) and FMT 4 (FR-conditioned, brown) into Il10^−/−^ mice. Mouse inflammation end-points are shown (triangles, red = active, green = inactive). Venn diagrams tally strains detected in inflamed (red), non-inflamed (green) Il10^−/−^ mice or found in both conditions (yellow) for **A** MP1 **B** MP2 **C** MP3 for each FMT and combined FMT 1, 3 and 4. **D-F** LEfSe (Linear discriminant analysis Effect Size) analysis of differentially abundant bacteria at species level based on metagenomic data in the recipient inflamed (active, red) or non-inflamed (inactive, green) Il10^−/−^ mice with abundance of strains in each condition. Different strains are labeled with different numbers. **D** MP1. **E** MP2. **F** MP3. **G** Venn diagrams shows shared or individual species (left) or strains (right) detected in MP1, MP2 and MP3. *zOTUs found in network analyses Fig 1G, S2.

Motivated by these differences, we next sought to identify species, strains, and metabolites that distinguished active *vs*. inactive *Il10*^−/−^ mice. Dozens of species showed significant differential abundance after the transfer experiments of MP1, MP2 and MP3 (40, 16 and 11, respectively **Fig. 5D-F**) including taxa from the network analyses and BONCAT experiments. Considering all microbiota transfer experiments, we detected 181 species, including 24 species shared between MP1-MP3 transfer experiments into *Il10*^−/−^ mice (**Fig. 5G, left** Vd). In contrast, none of the identified species shared any common strain within all (**Fig. 5G**, right Vd), demonstrating that active *vs*. inactive disease states in *Il10*^−/−^ mice are mediated by individually variable strain communities. Linear discriminant analysis Effect Size (LEfSe) determined 47 species that were differentially abundant between mice with active *vs*. inactive disease, each with individual strain repertoires in samples from the three different MPs (**Fig. S10A**). In the combined analyses *E. clostridioformis* (zOUT5) and *Bacteroides thetaiotaomicron* (zOUT2, zOTU66) were most dominant in mice with active disease and *Akkermansia muciniphila* (zOUT65) and *Escherichia coli* (zOTU4, zOUT8) were most dominant in mice with active disease (**Fig. S10A**). In addition, 56 species were significantly different in abundance comparing mice under chow vs EEN-like diet (**Fig. S10B**) with previously highlighted strains abundant in both groups, as mice colonized with *ex vivo* EEN-conditioned microbiota received either chow or EEN-like diet. Importance of strain level diversity, is obvious as several species were associated to both, active and inactive mice, in the different MP transfers, but the different strains were responding differently in the model systems (**Fig. 5D-G, S10A**).

SPLS-DA of untargeted metabolomics from mouse colon content separated active and inactive mice in the respective transfer experiments independent of dietary exposure (**Fig. S9B-D, S10C-F**). Several bile acids (BA) were highlighted as differentially abundant, which was confirmed in the targeted measurement primary taurine conjugated and secondary unconjugated BA (**Fig. S9B, C**). Targeted analyses of Isobutyrate showed highest concentration in mouse feces samples colonized with EEN conditioned microbiota on EEN-like diet (**Fig. S9E**). SPLS-DA of the combined untargeted metabolomics dataset confirmed distinct clusters for active and inactive disease independent of MP1-3 donor samples (**Fig. S10F**, **Fig. S11**), suggesting that individual strain profiles convey protective or aggressive responses in the IBD susceptible host (**Fig. S12**). These results clearly demonstrate that despite the model patients having personalized strain composition (**Fig. 5G**, right Vd), disease activity in gnotobiotic mice receiving FMTs was consistently associated with a number of metabolic and species-level taxonomic differences. Furthermore, the differentially abundant species in all humanized mice exhibited unique patterns linked to EEN and its cessation in the CD patient cohort (**Fig. S13**), emphasizing both the clinical relevance of our mouse FMT experiment findings and the individual microbial fingerprint of each patient.

In summary, we provide evidence that EEN impacts the individual gut microbiome – EEN therapy in the CD-EEN patients, selected fecal samples from three individuals exposed to MCFAs and EEN-like exposure in a non-host and host environment – leading to individual microbiota changes that are functionally relevant and harboring the capacity to induce or not to induce inflammation in mice. Despite the limited selection of donor samples, of the 16 EEN associated zOUTs from the network analyses, 7 zOTUs were found in significantly different abundant taxa in the mouse experiments (**Fig. 5D-F**) and 8 zOTUs were identified in the BONAT experiments (**Fig 2E, S5D**), suggesting a direct link of MCFAs in EEN to changes in the fecal microbiota of CD patients. The microbiota transfer studies in the gut chemostat model provides proof-of-concept that microbiota changes are functionally relevant and diet shapes individual microbiota signatures harboring the capacity to induce or not to induce inflammation in mice. In conclusion, we provide evidence that EEN therapy creates explicit functional changes of temporally and individually variable microbiome profiles in CD patients. These data challenge the concept that protective microbiome signatures are universal to patients and therapeutic interventions, instead we postulate individually diverse microbiome signatures with shared functional properties under these therapeutic conditions.

## Discussion

We followed 78 pediatric IBD patients longitudinally, focusing on 20 newly diagnosed CD patients receiving EEN as first-line treatment. Our results corroborate previous observations that EEN is highly effective at inducing remission. We demonstrate that protective functions of EEN therapy are mediated through diet-induced changes in patient microbiomes despite considerable variations in individual microbiota over time. Dietary exposure of EEN components like MCFAs and EEN-like diets on individual patient-derived fecal microbiota modified the activity and function of selective taxa. EEN mediated changes of fecal bacteria in the chemostat models convey anti-inflammatory phenotypes after FMT into IBD-relevant GF mice, strongly supporting a causal role of EEN-mediated protective changes in the microbiome. In contrast, fiber-repleted diets antagonized these protective functions. Notably and based on the model conditions, we identify patient-specific species and strain signatures in response to EEN therapy, confirming the highly personalized strain-level dynamics in patients and the absence of universal protective signatures.

Longitudinal sampling revealed an integrated network of taxonomic and metabolomic signatures across pediatric CD patients. In line with previous findings^20^, EEN therapy was associated with the genus *Enterocloster* (Lachnospiraceae; former genus *Lachnoclostridium)*. In addition, several *Enterocloster* species responded to MCFA exposure, *E. clostridioformis* (zOUT5) was the most abundant species in inactive *Il10*^−/−^ mice and showed high dynamics in the CD-EEN cohort together with *E. bolteae* (zOTU5). Low levels of *Lachnoclostridium* have been associated with CD^21^, supporting a protective function of *Enterocloster* in response to EEN therapy. Nevertheless, taxonomic shifts at the cessation of EEN were highly patient-specific. For example, while in the majority of patients, *E. bolteae* (zOTU5) was at much higher relative abundance during EEN compared to PostEEN, the dominant strains in many patients changed over this transition, and most strains were detected in just one subject. This individual diversity and turnover contribute to challenges in predicting taxonomic responses to dietary changes globally. Antibiotics and pre-existing dysbiosis are both associated with increased strain engraftment in patients receiving fecal microbiota transplants^22^. Likewise, the microbiome perturbation caused by EEN - and its cessation - may encourage strain turnover in some species, with potentially complex consequences for microbial function after therapy. Beyond taxonomic alterations, we highlighted six metabolites associated with EEN treatment and 16 metabolites associated with PostEEN samples. Independently, metagenomic analysis highlighted 583 bacterial gene functions with substantial changes in response to EEN, suggesting specific metabolic adaptations of the microbiota to host diets. Modulen IBD® is characterized by easily digestible carbohydrates and high fat content, including medium-chain triglycerides^9^. Our integrated network analysis highlighted caprylic (C8:0), decanoic (C10:0), and lauric (C12:0) acids as associated with EEN. MCFAs are part of the EEN formula and mediate anti-bacterial and anti-inflammatory functions^23–25^. The exposure of LA, DA and OA to selected patient samples directly link nutritional components of EEN to selectively activated taxa in the microbiota of CD patients. Surprisingly, from the limited selection of donor samples, 50% (8 out of 16 zOTUs) of the EEN associated bacteria significantly responded in the BONCAT experiments. Diets high in fat trigger release of bile acids^26^, providing a rationale for *C. innocuum* and *Hungatella hathewayi* to be associated with EEN ^27,28^. Concordantly, our untargeted metabolomics analysis identified several bile acids as differentially abundant, highlighting a potential microbial conversion of primary to secondary bile acids in non-inflamed *Il10*^−/−^ mice, potentially promoting wound healing and epithelial regeneration^26^. In a recent study (FARMM), EEN specifically affected amino acid pathways in the microbiome creating a metabolite milieu low in bacteria-derived indoles^17^, which was also low abundant in the EEN-like mouse diet. In addition, EEN-related protective effects in *Il10*^−/−^ mice are partially mediated by Isobutyrate production^16^, which was highest in mice receiving EEN-conditioned microbiota on EEN-like diet, supporting the hypothesis that microbiota and metabolite changes contribute to the EEN therapeutic efficacy.

In healthy humans, dietary fiber intake supports beneficial functions of the gastrointestinal tract^29^. In animal models, low fiber diets induce adverse microbiota changes associated with impaired intestinal barrier integrity and bacteria-related mucus degradation^30,31^, highlighting the fiber paradox in understanding EEN-mediated anti-inflammatory mechanisms in IBD. Dietary fiber consists of complex carbohydrates, which are resistant to digestion and absorption by the host. However, dietary fiber degradation and availability for the host depends on the fermenting potential of host-specific microbial communities, which are modified by intestinal inflammation^29,32–34^. Using the combined gut chemostat-to-mouse transfer approach, we showed that a fiber-free EEN-like medium maintains the anti-inflammatory and remitting phenotype associated with EEN therapy. Concordantly, the protective effect of EEN-conditioned microbiota is reversible upon exposure to fiber-repleted diet, as a proxy for regular diet PreEEN and PostEEN, and potentially resistant to repetitive EEN as shown before^12^. A potentially adverse effect of fiber in IBD was elucidated by Armstrong and colleagues, demonstrating the incapability and loss of function of bacteria to degrade fibers promoting intestinal inflammation^34^. Furthermore, several additional studies link the presence of dietary fiber with increased inflammation^35,36^. In addition, fiber-free diets prevent and diminish the colonization of identified pathobionts in different CD-like mouse models^37,38^, supporting our hypothesis that EEN therapy directly creates protective niches in the gut microbiome. Preserving a beneficial microbiome after cessation of EEN may be a new approach to guide mucosal healing and remission in pediatric CD^39^. In our IBD cohort, 40% (8/20) of pediatric CD patients relapsed within the first year after EEN treatment, highlighting the necessity to maintain remitting conditions. Our data clearly demonstrate that EEN therapy mediates protective changes in individually variant microbiome profiles, leading towards personalized patient-to-patient species and strain signatures. These findings support the idea to develop patient-tailored maintenance strategies in pediatric CD involving combinations of personalized nutritional and bacterial interventions.

## Acknowledgement

We thank the patients and their families for participating in our research.

## Author contribution

DHaller and TS are responsible for the conceptual design of the study; DHäcker and DHaller designed the *ex vivo* experiments; KSiebert, HHölz, JHeetmeyer, FDeZen, GLeThi, TS: collected and prepared clinical data and samples, accompanied, followed and characterized patients, analysis of clinical data; HHölz, TS recruited patients; DHäcker, HHeimes, AMetwaly and DHaller performed and interpreted mouse and fermenter experiments; AR and JN performed BONCAT experiments and analyses; DHäcker performed 16S rRNA gene amplicon and corresponding statistical analysis with support of QM and ML; CM, QM, KSocas, ML, KK performed metabolomics with statistical analysis; KSiebert, DHäcker, NK, AMetwaly, ML responsible for integration of clinical metadata with 16S and metabolomics as well as sample selection and performance of data integration and network analysis; NK, ML, PJK performed 16S & untargeted metabolomics data integration, untargeted metabolomics data analyses; BS, AMahapatra, KSP performed metagenomic sequencing and analyses; KN provided 16S rRNA gene amplicon and metagenome sequencing and support for analysis; DHäcker, KSiebert, BS, NK, KSP, TS and DHaller evaluated and interpreted data and wrote the manuscript; All authors provided input on the manuscript, critically revised and approved the final version of the manuscript.

## STAR Methods

### RESOURCE AVAILABILITY

#### Lead contact

Further information and requests should be directed to and will be fulfilled by the lead contact Prof. Dr. Dirk Haller.

#### Materials availability

This study did not generate new unique reagents.

#### Data and code availability

##### Data availability

- Some data that support the findings of this study will be available on reasonable request from the corresponding author [TS]. The data are not publicly available due to the pediatric age of the research participants.
- Mouse untargeted metabolomics data is available at Massive (https://massive.ucsd.edu) under the number MSV000094047.
- 16S rRNA gene sequencing results of the cohort, mouse and fermenter experiments will be available at the time of publication.

##### Code availability

- Mutliomic data integration of 16S rRNA and metabolomics code is available at https://doi.org/10.5281/zenodo.10808412.
- All code necessary to reproduce the metagenomic data analysis will be made publicly available at the time of publication

### Experimental model and study participant details

#### Patient cohort, recruitment and study design

Patients (aged 3-18 years) with suspected or established IBD were recruited to a monocentric pediatric IBD cohort study (ethics approval Ludwig-Maximilians University of Munich, approval No. 17-801 and German Clinical Trials Register Accession No. DRKS00013306, date registration 19.03.2018). All patients and parents/legal guardians provided written informed consent/assent. Patients without confirmed IBD after full diagnostic work-up according to revised Porto criteria were excluded^40^. All other patients were followed prospectively for one year and treated according to current ESPGHAN/ECCO guidelines for CD or ulcerative colitis (UC)/IBD unclassified (IBDU)^6,41,42^. Disease activity, as well as endoscopic, histologic, radiologic and laboratory findings were collected prospectively. Disease activity indices included the Physician Global Assessment (PGA) and the weighted pediatric Crohn’s disease activity index (wPCDAI) for CD, as well as the pediatric ulcerative colitis activity index (PUCAI) for UC/IBDU. For EEN-treated CD patients, remission was defined as wPCDAI of <12.5 points and response as reduction in wPCDAI of >17.5 points, but not reaching remission. Relapse was defined as increase of wPCDAI to ≥12.5 points and/or fecal calprotectin of >250 mg/L requiring changes in medical treatment or abdominal surgery or reintroduction of EEN. Patients received detailed instructions for fecal specimen collections and time points, as well as stool collection kits. Each sample was accompanied with a questionnaire on date and time of specimen collection, problems during sampling, Bristol stool scale and recent antibiotic use.

EEN therapy was prescribed for 6-8 weeks, followed by gradual re-introduction of solid food and concomitant start of maintenance treatment. EEN was performed with Modulen® IBD (Nestlé Health Science) or in case of intolerance (e.g. cow’s milk protein allergy) with Neocate Junior® (Nutricia, Erlangen, Germany). The amount of prescribed formula was based on the Daily Recommendations Intake (DRI) using the online calculator: https://www.nal.usda.gov/human-nutrition-and-food-safety/dri-calculator. All EEN-treated patients received dietary advice and dietetic support to comply with complete liquid formula feeding.

#### Animal ethics statement

All animal experiments were conducted at Technical University of Munich and approved by the Committee on Animal Health Care and Use of the State of Upper Bavaria (TVA ROB-55.2-2532.Vet_02-19-190) and performed in strict compliance with the EEC recommendations for the care and use of laboratory animals (European Communities Council Directive of November 24, 1986 (86/609/EEC).

#### Animals and housing conditions

GF wild-type (WT) and *Il10*-deficient (*Il10*^−/−^) mice on 129Sv/Ev background were bred and kept at the Core Facility Gnotobiology of the ZIEL Institute for Food & Health, Technical University of Munich, Freising, Germany. Mice were housed as described elsewhere^19^, but using single gnoto-cage units (Isocage P System, Tecniplast, Italy). Cages are ventilated with HEPA-filtered air at 22 ± 1°C with a 12-h light/dark cycle. Female and male mice were assigned randomly to the treatment groups with a ratio of 1:1 and six mice per genotype. Mice received standard chow (V1124-300; Ssniff, Soest, Germany; autoclaved) or an EEN-like purified diet (EEN-like, SNIFF S5745-E902, **Fig. S14D**) and sterile water *ad libitum*. EEN-like diet was introduced one week prior to fecal microbial transfer (FMT) to allow for adaptation. Handling of mice in isocages was performed under biological safety cabinets in sterile conditions.

### Method details

#### Fecal specimen collection

Samples were collected at home or in the hospital. Patients provided samples at baseline (before bowel preparation for endoscopy) and every four weeks in (1) 4 mL DNA stabilizer (magiX PBI Microbiome Preservation Buffer, microBIOMix GmbH, Regensburg)^43^, (2) 8 mL 20% glycerol in reduced phosphate-buffered saline (PBS, 0.05% L-cysteine) and (3) naïve without stabilizing solution. Patients on EEN collected stool in (1) weekly for the first 12 weeks. Samples in (1) were shipped at ambient temperature by mail to the study center. Fecal specimens in (2) and (3) were immediately frozen at home (approx. −18 to −20°C) and brought in on wet ice packs at the next visit. Upon receipt, samples were aliquoted, barcoded, linked to clinical data with CentraXX Bio (Kairos GmbH) and stored immediately at −80°C until further analysis.

#### Selection of donor samples for BONCAT and transfer experiments into germ-free mice

We selected Stool samples for direct exposure of MCFAs and FMT experiments into germ-free (GF) mice to show proof-of-concept examples. Therefore, we selected samples based on sampling timepoint (include various states along EEN therapy: active disease sample from treatment-naive patient before the start of EEN (PreEEN, pE), EEN-induced remission (EEN, EE) and a relapse sample after reintroduction of a regular diet (PostEEN, PE)); clinical activity (pga, calprotectin), drug exposure (shorter time under co-medication preferred) and histopathology score in Il10^−/−^ mice of FMT1 (direct transfer). For MP1 (model patient 1) the pE was selected as EE and PE samples induced inflammation in the mouse model, so lowered disease activity in the patient seemed to be influenced by Anit-TNF. PreEEN sample form MP2 had very bad quality due to its bloody and diarrhetic consistency, therefore EE and relapse sample were chosen. PE sample in remission was not chosen as a direct impact of EEN therapy was no longer expected. EE sample from MP3 was chosen to investigate a sample where mucosal healing was not fully achieved during EEN therapy. Details are in **Table S3**.

#### Preparing gavage for humanizing germ-free mice

Preparation of FMT with human fecal microbiota was performed according to Metwaly *et al*. (2020)^19^. For post *ex vivo* FMT, frozen samples in glycerol (20% end concentration) were thawed on ice. All steps were performed in an anaerobic chamber (90% N_2_ and 10% H_2_). *Ex vivo* suspensions (2 mL) were centrifuged at 100×*g* for 3 min at 4°C to pellet debris. Supernatant was collected and centrifuged at 8000×*g* for 10 min at 4°C to pellet the bacteria. The bacterial pellet was resuspended in 1.5 mL reduced PBS (0.05% L-cysteine-HCl). The clear supernatant was transferred into an anaerobic crimped tube, which was transferred to the gnotobiotic facility. All mice were gavaged on three consecutive days and sacrificed after four weeks of colonization (12 weeks of age).

#### Tissue Staining and scoring

Colonic and cecal Swiss-roll tissues were fixed, stained and scored as described before^19^. In brief, tissue was fixed in 4% formaldehyde/PBS for 48 h at room temperature, subsequently dehydrated (Leica TP1020), and embedded in paraffin (McCormick; Leica EG1150C). Formalin-fixed paraffin-embedded (FFPE) tissue sections were stained with hematoxylin and eosin (H&E) and scored 0 (not inflamed) to 12 (highly inflamed). Inflammation cut off was defined as a histopathology score greater than three based on scoring in WT mice.

#### Gene expression

Tissue sections from colon were collected in RNAlater (Sigma Aldrich) for 24 h at 4°C and long-term at −80 °C until processing. Total RNA from tissue was isolated by using the NucleoSpin RNA II kit (Macherey-Nagel GmbH) according to manufacturer’s instructions. Complementary DNA was synthetized from 500 ng total RNA by using random hexamers and moloney murine leukemia virus (M-MLV) reverse transcriptase (RT) Point Mutant Synthesis System (Promega). Quantification was performed by using the LightCycler 480 Universal Probe Library System (Roche Primers and probes used were: *Gapdh* (glyceraldehyde 3-phosphate dehydrogenase) (5’-tccactcatggcaaattcaa-3′; 5’ tttgatgttagtggggtctcg-3′; Probe #9) and *Tnf* (Tumor necrosis factor) (5′-tgcctatgtctcagcctcttc 3′; 5′-gaggccatttgggaacttct-3′; Probe #49). Relative induction of gene mRNA expression was calculated based the 2-ΔΔCt method ^44^ using the expression of *Gapdh* for normalization.

#### *Ex vivo* gut chemostat model

The *ex vivo* continuous fermentation was performed in 1.4-L bioreactors (Multifors 2) and monitored with Eve®software (Infors AG, Basel, Switzerland). Parameters were set as described before^45^. Briefly, vessels were maintained at 37°C, pH 6.8 with a constant influx of N_2_ to maintain an anaerobic atmosphere. Inoculation material was prepared in an anaerobic chamber (90% N_2_ and 10% H_2_). Approximately 2.5 g of frozen fecal samples was suspended in reduced PBS supplemented with 0.03 g/mL L-cysteine-HCl, filtered (70 µm Cell Strainer, FisherScientific) and evenly distributed between two vessels, which led to about 1.0×10^8^ to 8.16×10^8^ CFU/vessel. Two consecutive batch fermentations (24 h in 400 mL, another 24 h with fresh 400 mL medium added) allowed fecal microbial communities to establish. Continuous fermentation included complete medium exchange every 36 h (retention time) with a working volume of 400 mL. After 10 days of continuous fermentation, where stable community formation was achieved, samples for transfer into GF mice were collected and medium was switched afterwards. After 1.5 days of complete medium exchange from medium 1 to medium 2, samples for transfer into mice were collected on day 9 of continuous fermentation in the second medium. Medium composition was either fiber-rich (FR) or EEN-like, mimicking patient conditions at sampling (PreEEN and PostEEN, respectively; **Fig. S8A-E**).

FR medium was prepared according to Macfarlane *et al*. (1998)^46^, but supplemented with additional vitamins (pantothenate 10 mg/L, nicotinamide 5 mg/L, thiamine 4 mg/L, biotin 2 mg/L, vitamin B12 0.5 mg/L, menadione 1 mg/L, and p-aminobenzoic acid 5 mg/L) as described in Gibson and Wang^47^. EEN-like medium was based on the Macfarlane medium, but with modifications simulating colonic-luminal content during an EEN diet. To determine the carbohydrate:protein ratio of digested Modulen IBD® in the colon, similar calculations as described previously were conducted^48,49^. Since Modulen IBD® lacks fibers, these ingredients present in the FR medium (i.e., pectin from citrus, guar gum, xylan from oat spelt, inulin from *Dahlia* tubers, and arabinogalactan from larch wood) were replaced by soluble rice starch and simple sugars. A total carbohydrate concentration of 13 g/L was chosen in alignment with Cinquin *et al.* (2004)^48^. As the formulation of Modulen IBD® is not publicly available, we decided to include some Modulen IBD® not to miss crucial nutrients in our model-colon medium. Towards this end, 10% of the 1kcal/mL standard preparation of Modulen IBD® was added to 1L of EEN-like medium. Finally, the carbohydrate:protein ratio was 75:25 in the model medium. To further mimic Modulen IBD®, casein was used as main protein source with the exclusion of tryptone and peptone (**Fig. S14A-C**).

Samples were aseptically collected daily at the same daytime and stored at −80°C until further analyses. Samples for transfer into the GF mice were collected and mixed with glycerol (20% end concentration) and stored at −80°C until gavage preparation. In order to diminish potential vessel effects in the technical duplicates of each fermenter colonization, samples from both vessels were mixed 50:50 for gavage.

#### High-throughput 16S-ribosomal RNA (rRNA) gene amplicon sequencing analysis

Total DNA from human stool, colonic mouse content and the continuous *ex vivo* fermentation was isolated using bead beating and sequenced for 16S rRNA gene amplicons as described^50^. Downstream analysis was performed in R v4.2.1 using Rhea (https://lagkouvardos.github.io/Rhea/)^51^. zOTU tables were normalized to 10.000 reads, beta-diversity was computed using generalized UniFrac distances^52^. Alpha-diversity was assessed on the basis of taxa richness and Shannon effective number of zOTUs^53^. Trees were visualized and annotated using EvolView (http://www.evolgenius.info/evolview/)^54^. Differential analyses of bacteria in different groups was done using LDA Effect Size (LEfSe)^55^. zOTUs were re-checked using the 16S-based ID tool of EzBioCloud^56^ using the database update 2023.08.23, reflecting all taxa published and accepted by IJSEM until April 2023.

#### Shotgun Metagenomic Sequencing and Analysis

Metagenomic sequencing was performed for 96 human stool samples and 66 samples from transfer experiments (54 mouse, 12 *ex vivo*) using the genomic DNA already isolated for 16S rRNA gene amplicon sequencing. DNA was randomly sheared into short fragments. The obtained fragments were end repaired, A-tailed, and further ligated with Illumina adapters. The fragments with adapters were size-selected, PCR amplified, and purified. The library was checked with Qubit and real-time PCR for quantification and BioAnalyzer for size distribution detection as required by Illumina (TruSeq® DNA Library Prep Kit). Quantified libraries were pooled and sequenced on a NovaSeq platform (Illumina) in PE150 mode. Raw data were aimed at 15 Gb per demultiplexed sample. From raw shotgun metagenomic sequencing data, adapter sequences were removed using Trimmomatic v0.39^57^ without considering any mismatch and parameters LEADING:3, TRAILING:3, and SLIDINGWINDOW:4:15. We removed human reads by mapping against human reference genome hg38 by using Bowtie2 v2.4.5 ^58^ with the default parameters. Cleaned fastq data files were used in the following steps.

Strain tracking and strain gene content analyses were performed based on the StrainPGC workflow^59^. Briefly, preprocessed metagenomes were metagenotyped using GT-Pro v1.0.1^60^. Across all samples, 369 species - as defined in the Unified Human Gut Genome (UHGG)^61^ - were robustly detected and only these were included in downstream pangenome profiling. Single nucleotide polymorphism (SNP) profiles were filtered, subsampled, and strain relative abundances were estimated with StrainFacts v0.4.0^18^. Within individual species, estimated strain fractions add up to 100%. By contrast, other analyses considered the overall strain composition: the relative abundance of each strain across all species.

Gene profiling was performed against a subset of the MIDASDB-UHGG pangenome database v.15^62^ dereplicated at a 99% ANI threshold performed and mapping reads using Bowtie2 v2.4.5^58^. Species abundances were estimated based on the average depth of core genes. Global functional gene family profiling was performed by aggregating gene depths into Clusters of Orthologous Genes (COGs) across all species and then normalizing based on 25 ubiquitous, single-copy genes to estimate “gene copies per genome”.

#### *Ex vivo* anaerobic incubations with medium chain fatty acids (MCFAs)

Fecal samples (MP1 PreEEN, MP2 EEN, MP2 relapse, and MP3 EEN) previously collected in 20% glycerol in reduced phosphate-buffered saline (PBS, 0.05% L-cysteine) were thawed and introduced into an anaerobic chamber (10% H_2_, 90% N_2_). All reagents and materials were introduced in the anaerobic chamber 48 hours before sample processing to ensure anaerobic state by the start of the experiment.

Stool samples (1-1,5 g) were homogenized with 1X PBS (1:10 dilution) and filtered using a 40 μm size filter (Corning, Germany) to remove large particles, which was followed by three washing steps in 1X sterile PBS to remove residual of glycerol and leftover nutrients in the stool. Afterwards, samples were added to sterile falcon tubes containing a final concentration of 50µM of the cellular activity marker L-azidohomoalanine (AHA), (Baseclick GmbH, Germany) and 10µM of MCFAs (lauric acid, decanoid acid and octanoic acid) dissolved in ethanol with a final concentration of 0.5%. 0.5% ethanol was used as negative control and 2 mg/mL of glucose as positive control for each experiment. Samples were incubated for 24 hours under anaerobic conditions. After 24 hours of incubation, part of the samples were frozen for DNA extraction and other aliquots were washed twice in PBS and then fixed in 1:1 ethanol:PBS for further analysis with fluorescence activated cell sorting (FACS). Both ethanol fixed and frozen samples were collected in triplicates^63,64^.

#### Bioorthogonal non-canonical amino acid tagging (BONCAT) and images analysis of translationally active bacteria

To identify utilization capability of MCFAs by the gut microbiota, we employed BONCAT, a fluorescence-based single cell labeling technique. BONCAT involves incorporation of the non-canonical amino acid AHA to replace L-methionine during protein synthesis, followed by fluorescent labeling of AHA-containing proteins using azide-alkyne click chemistry^65^ (**Fig. 2A**). Cu(I)-catalyzed click chemistry reaction was performed on microscopy slides as previously described^65^ and counter-stained with 4′, 6-diamidino-2-phenylindole (DAPI).

20 pictures for each sample were collected using a confocal microscope (Olympus, fluoview, FV10i, Germany) and processed using Fiji^66^. Translationally active cells were quantified using the software *digital image analysis in microbial ecology* (Daime)^67^. Translationally active cells were measured as biovolume fraction comparing the Cy5-labeled cells (BONCAT positive) to the total biomass stained with DAPI.

#### BONCAT-FACS and DNA extraction

A combination of BONCAT and FACS was used to determine which bacteria are capable of MCFAs utilization. Briefly, ethanol fixed samples were washed in 1X sterile PBS, resuspended in 96% ethanol and centrifuged again. Afterwards, the BONCAT dye solution was added to the samples and incubated for 30 min in the dark at room temperature^65^. After incubation, samples were washed three time in 1X sterile PBS and filtered with a 35 mm nylon mesh using BD tubes 12x 75 mm (Corning, Germany) right before sorting. Cy5-positive bacteria were sorted, collected into sterile tubes and store at-80°C until DNA extraction. A representation of the gating strategy is shown in Supplementary Figures 1.

Bacterial DNA from sorted and unsorted samples were extracted using the QIAmp DNA mini kit (Qiagen, Germany) following the protocols for bacteria according to the manufacturer’s instructions with an additional lysozyme treatment^64^. Every sample was sorted in triplicates. FACS data were further analyzed with FlowJo™ v10.10.0 Software (BD Life Sciences; Reference: FlowJo™ Software (for Windows) Version v10.10.0. Ashland, OR: Becton, Dickinson and Company; 2023).

#### BONCAT data analysis

Statistical analysis was performed using R statistical software (https://www.r-project.org/). 16S rRNA amplicon sequencing analysis was carried out with Rhea, beta diversity was assessed using UniFrac distances as previously described ^51,52^ and the statistical significance of factors was evaluated using permutational multivariate analysis of variance (perMANOVA). Differences in relative abundance between sorted and unsorted fractions and between sorted MCFAs and ethanol control, were analyzed using ANOVA and Tukey’s test for multiple comparisons. For quantifications of the percentage of active cells, Kruskal-Wallis and Dunn’s test were applied. Significantly enriched and depleted zOTUs sequences were identified using the 16S-based ID tool of EzBioCloud^56^.

#### Metabolomics

##### Sample preparation / Metabolites extraction

###### Sample preparation of mouse colon content

Approximately 20 mg of mouse colon content was weighed in a 2 mL bead beater tube (FastPrep-Tubes, Matrix D, MP Biomedicals Germany GmbH, Eschwege, Germany). Next, 1 mL of methanol-based dehydrocholic acid extraction solvent (c = 1.3 µmol/L) was added as an internal standard for work-up losses. The samples were extracted with a bead beater FastPep-24TM 5G (MP Biomedicals Germany GmbH, Eschwege, Germany) supplied with a CoolPrepTM (MP Biomedicals Germany, cooled with dry ice) 3-times each for 20 s of beating at a speed of 6 m/s and followed by a break of 30 s each.

###### Sample preparation of fermenter or human samples

Either 200 mg of the fermenter samples or 100 mg of human fecal samples was weighed in a 15 mL bead beater tube (FastPrep-Tubes, Matrix D, MP Biomedicals Germany GmbH, Eschwege, Germany). Next, 5 mL of methanol-based dehydrocholic acid extraction solvent (c = 1.3 µmol/L) was added as an internal standard for work-up losses. The samples were extracted with the same FastPrep protocol described above.

###### Targeted bile acid measurement

A standard of 20 µL of isotopically labeled bile acids (ca. 7 µM each) were added to 100 µL of sample extract. Targeted bile acid measurement was performed using a QTRAP 5500 triple quadrupole mass spectrometer (Sciex, Darmstadt, Germany) coupled to an ExionLC AD (Sciex, Darmstadt, Germany) ultrahigh performance liquid chromatography system. A multiple reaction monitoring (MRM) method was used for the detection and quantification of the bile acids according to Reiter et. al.^68^ Data acquisition and instrumental control were performed with Analyst v1.7 software (Sciex, Darmstadt, Germany).

###### Untargeted mass spectrometric measurement

The untargeted analysis was performed using a Nexera UHPLC system (Shimadzu, Duisburg, Germany) coupled to a Q-TOF mass spectrometer (TripleTOF 6600, AB Sciex, Darmstadt, Germany). Separation of the fecal samples was performed either using a UPLC BEH Amide 2.1 × 100 mm, 1.7 µm analytic column (Waters, Eschborn, Germany) with a 400 µL/min flow rate or with a Kinetex XB18 2.1 × 100 mm, 1.7 µm (Phenomenex, Aschaffenburg, Germany) with a 300 µL/min flow rate. For the HILIC-separation the settings were as follows: The mobile phase was 5 mM ammonium acetate in water (eluent A) and 5 mM ammonium acetate in acetonitrile/water (95/5, v/v) (eluent B). The gradient profile was 100% B from 0 to 1.5 min, 60% B at 8 min and 20% B at 10 min to 11.5 min and 100% B at 12 to 15 min. For the reversed-phase separation, eluent A was 0.1% formic acid and eluent B was 0.1% formic acid in acetonitrile. The gradient profile started with 0.2% B which was held for 0.5 min. Afterwards, the concentration of eluent B was increased to 100% until 10 min which was held for 3.25 min. Subsequently, the column was equilibrated at starting conditions. A volume of 5 µL per sample was injected. The autosampler was cooled to 10 °C and the column oven heated to 40 °C. Every tenth run, a quality control (QC) sample, which was pooled from all samples, was injected. The samples were measured in a randomized order and in the Information Dependent Acquisition (IDA) mode. MS settings in the positive mode were as follows: Gas 1 55, Gas 2 65, Curtain gas 35, Temperature 500 °C, Ion Spray Voltage 5500, declustering potential 80. The mass range of the TOF MS and MS/MS scans were 50–2000 m/z and the collision energy was ramped from 15–55 V. MS settings in the negative mode were as follows: Gas 1 55, Gas 2 65, Cur 35, Temperature 500 °C, Ion Spray Voltage −4500, declustering potential −80. The mass range of the TOF MS and MS/MS scans were 50–2000 m/z and the collision energy was ramped from −15–55 V.

##### Data processing

The “msconvert” from ProteoWizard^69^ was used to convert raw files to mzXML (de-noised by centroid peaks). The bioconductor/R package xcms^70^ was used for data processing and feature identification. More specifically, the matchedFilter algorithm was used to identify peaks (full width at half-maximum set to 7.5 s). Then the peaks were grouped into features using the “peak density” method^70^. The area under the peak was integrated to represent the abundance of features. The retention time was adjusted based on the peak groups presented in most samples. To annotate features with names of metabolites, the exact mass and MS2 fragmentation pattern of the measured features were compared to the records in HMBD^71^ and the public MS/MS spectra in MSDIAL^72^, referred to as MS1 and MS2 annotation, respectively. Missing values were imputed with half of the limit of detection (LOD) methods, i.e., for every feature, the missing values were replaced with half of the minimal measured value of that feature in all measurements. To confirm a MS2 spectra is well annotated, we manually reviewed our MS2 fragmentation pattern and compared it with records in the public database, previously measured reference standards and SIRIUS^73^ to evaluate the correctness of the annotation. Significances of well-annotated metabolic features were computed with analysis of variance (ANOVA) per donor or Wilcoxon signed-rank tests and FDR-corrected using the Benjamin-Hochberg method for three donors. Subsequent hierarchical clustering using only differentially abundant metabolites (q < 0.05) was conducted using complete linkage based on Euclidean distances. Calculations, as well as heatmaps, were computed in R v4.2.1. For each principal component analysis (PCA), we included negative and positive features from RP and HILIC with at least 50% non-missing values per condition (inflamed/non-inflamed). The remaining missing values were imputed as described above. The prcomp function from R v4.3.2 computed the principal components after centering and scaling each feature to zero mean and unit variance. Sparse Partial Least Squares Discriminant Analysis (sPLS-DA) was performed on centered and scaled features using the mixOmics bioconductor/R package v6.26.0(Singh et al., 2019). Mean balanced error rate, was used to select the number of components and features in M-fold cross-validation where M corresponds to the minimum number of samples per condition. **Fig. S11** shows raw values for features with the highest Variable Importance in the Projection (VIP), i.e., features contributing the most to explaining the conditions.

##### Data integration of 16s rRNA amplicon data with metabolomics and metagenomics

We matched 60 samples from 15 patients of the 16S-rRNA gene sequencing to 60 samples of the untargeted metabolomics analysis according to patient ID and same time points. The aim was to select features that jointly discriminate the 23 EEN samples (clinical phenotype mild and remission) and 37 PostEEN samples (>2 weeks after EEN, clinical remission). For the integrated selection of metabolome and microbiota features, sPLS-DA (mixOmics v6.20) was used(Singh et al., 2019). For feature selection concerning metabolomics, only annotated metabolites were considered. As described above, input metabolites were mean-centered and scaled to unit variance. zOTUs (zero-radius operational taxonomic units) were centered log-ratio transformed. To estimate the degree of overfitting, the model was trained on 70% of the data, with a k-fold cross-validation for feature selection during training. The remaining 30% of the data were used to evaluate the generalization power. The model performance was measured with an Area Under the Receiver Operating Characteristic (ROC-AUC) on the samples in the test set. Subsequently, Spearman’s rank correlation coefficients between all selected metabolites and zOTUs were computed and their respective p-values were corrected with the Benjamini-Hochberg procedure^75^. All coefficients with an FDR >0.05 were set to 0. Using all non-zero correlations, a network was generated and the Louvain method for community detection^76^ was applied to extract highly connected subgraphs from the correlation network. For each community, a sPLS-DA model was trained with the same train-test scheme explained previously for feature selection. Using the mean loadings scores over all k iterations, the most relevant metabolites and zOTUs were identified. More precisely, for metabolites and zOTUs separately, the absolute mean loadings were plotted in ascending order and all features on the right of the knee point (20 zOTUs and 22 metabolites) were selected. Analyses were run in R v4.2.1 and python v3.8.11. For network analysis the igraph v1.3.4^77^ and networkx v3.1^78^ packages were used.

Profiles of 16S rRNA amplicon relative abundance were further integrated with metagenomic species profiles in order to identify those which represent the same, underlying taxa. This was done by cross-decomposition of zOTU relative abundance and species relative abundances estimated from the metagenomic workflow, using the PLSSVD model implemented in SciKit-Learn^79^, multiplying the matrices of weights for each data-type provides a matrix of coefficients mapping zOTUs to metagenomic species. We scaled these coefficients by the square-root of the mean relative abundance of that zOTU and species, respectively, producing a matrix of matching scores for each comparison, while accounting for all other taxa. Based on this score, we ranked zOTUs for each species and species for each zOTU. We selected as putative matches all zOTU-species pairs that were either reciprocal best hits or a combination of one best hit and one second-best hit, and where at least one of the genus or family classifications matched. In this way, we accounted for the possibility of multiple zOTUs representing a species or multiple species being represented by a zOTU.

##### Statistical analysis

MDS and PCA latent dimensions in **Fig. 1** and **Fig. S1** were tested for significance using linear mixed models of the form

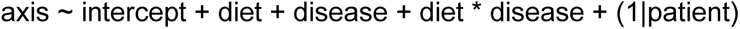

where diet, disease and their interaction are included as fixed effects and patients are included as random effects. All coefficients are tested with a t-test for **Fig. 1** and an ANOVA for **Fig. S1**. Reported p-values are Bonferroni-corrected. To estimate the degrees of freedom, we used Satterthwaite’s method as implemented in the lmerTest package (version 3.1-3). Further statistical analyses and plots were generated using GraphPad Prism v8.0 (GraphPad, La Jolla, CA) using Mann-Whitney test, ANOVA followed by pairwise comparison testing (Tukey, Bonferroni correction), by Kruskal-Wallis test followed by Dunn’s multiple comparisons. Data is presented as mean ± SD. P-values below 0.05 were considered significant (p < 0.05, *; p < 0.01, **; p < 0.001, ***; and p < 0.0001, ****).

Taxonomic turnover in both zOTU and strain compositions were calculated based on the Bray-Curtis dissimilarity. Pairs where one sample was collected during EEN and the other during PostEEN were classified as “transition”. All within-subject pairwise comparisons were computed and fit using cubic spline regression of the time between sample collections, with a regression term for each subject, as well as for each pair class: EEN, transition, or PostEEN. The null hypothesis of no difference between pair classes was tested by permutation: 999 random permutations of samples within subjects. Changes in mean, normalized, COG depths during EEN and PostEEN time points were tested using the Wilcoxon signed-rank test across subjects. False discovery rates were calculated using the Benjamini-Hochberg procedure. Mean fold change for each COG was calculated as the ratio between these two estimates.

### Additional resources

#### Clinical Trial Registry

The study is registered at the German Clinical Trials Registry under the Accession No. DRKS00013306, date of registration 19.03.2018.

## Supplementary Data

### Supplemental information

**Document S1. Figures S1-S14, Tables S1-S5:**

Figures S1-S3 related to Figure 1.

Tables S1-S2 related to Figure 1.

Table S3 related to donor sample selection related to Figure 2-5

Figures S4-S5 and tables S4-S5 related to Figure 2

Figures S6 and tables S6 and S7 are related to Figure 3.

Figures S7-S8 related to Figure 4.

Figures S9-S13 related to Figure 5.

Figures S14 related to media and diet information in STARMethods.

Tables S6 and S7: Excel files containing additional data too large to fit in a PDF. Related to Figure 3.

