## Supplemental Material for "Exclusive Enteral Nutrition Initiates Individual Protective Microbiome Changes to Induce Remission in Pediatric Crohn’s Disease"

**Supplemental information**

Figures S1-S3 related to Figure 1.

Tables S1- S2 related to Figure 1.

Tables S1 related to donor sample selection related to Figure 2 -5

Figures S4-S5 and tables S4-S5 related to Figure 2

Figures S6 and tables S6 and S7 are related to Figure 3. Tables S6 and S7: Excel files containing additional data too large to fit in a PDF.

Figures S7-S8 related to Figure 4.

Figures S9-S13 related to Figure 5.

Figures S14 related to media and diet information in STARMethods.

| | | CD (N = 45) | CD-EEN (N = 20) | | UC /IBDU (N = 33, 29/4) |
|---|---|---|---|---|---|
| Sex | Male | 24 (53%) | 11 (55%) | | 18 (55%) |
| | Female | 21 (47%) | 9 (45%) | | 15 (45%) |
| Age at diagnosis (years) | | 11.9 (3.5) | 12.4 (3.2) | | 9.6 (4.3) |
| Age at study inclusion (years) | | 13.3 (3.3) | 12.5 (3.1) | | 10.8 (4.2) |
| First degree family history of IBD | | 8 (18%) | 3 (15%) | | 7 (21%) |
| Mean height (z-score) | | -0.06 (1.05) | 0.17 (1.06) | | 0.24 (1.12) |
| Mean weight (z-score) | | -0.51 (1.29) | -0.57 (1.32) | | -0.23 (1.08) |
| Mean body-mass index (z-score) | | -0.75 (1.49) | -0.95 (1.44) | | -0.57 (1.03) |
| **Disease characteristics** | | | | | |
| Location - lower GI [1] | None | 1 (2%) | 1 (5%) | Proctitis | 0 |
| | Ileum only | 8 (18%) | 3 (15%) | Left sided | 4 (12%) |
| | Colon only | 8 (18%) | 5 (25%) | Extensive | 7 (21%) |
| | Ileocolonic | 28 (62%) | 11 (55%) | Pancolitis | 22 (67%) |
| Location - upper GI [1] | Proximal (n, %) | 34 (76%) | 18 (90%) | | |
| | Distal (n, %) | 11 (24%) | 6 (30%) | | |
| Behavior [1] | Nonstricturing-nonpenetrating | 37 (82%) | 17 (85%) | Never severe | 23 (70%) |
| | Stricturing | 5 (11%) | 2 (10%) | Ever severe | 10 (30%) |
| | Penetrating | 1 (2%) | 1 (5%) | | |
| | Stricturing and penetrating | 2 (4%) | 0 | | |
| Perianal involvement | | 9 (20%) | 4 (20%) | | |
| Evidence of growth delay | | 11 (24%) | 4 (20%) | | |
| Extraintestinal manifestations [2] | | 8 (18%) | 5 (25%) | | 5 (15%) |
| Previous surgery | | 5 (11%) | 0 | | 0 |
| **Disease activity** | | | | | |
| Median wPCDAI | | 40 (30 to 52.5) | 43.8 (38.1 to 51.3) | PUCAI | 40 (25 to 57.5) |
| Inactive (wPCDAI <12.5) | | 3 (7%) | 0 (%) | Inactive (PUCAI <10) | 3 (10%) |
| Mild (wPCDAI 12.5 to 40) | | 23 (51%) | 8 (40%) | Mild (PUCAI 10 to 34) | 11 (33%) |
| Moderate (wPCDAI 42.5 to 57.5) | | 11 (24%) | 8 (40%) | Moderate (PUCAI 35 to 64) | 12 (36%) |
| Severe (wPCDAI >57.5) | | 8 (18%) | 4 (20%) | Severe (PUCAI ≥65) | 7 (21%) |
| **Baseline laboratory data** | | | | | |
| Mean CRP (mg/dL) | | 2.7 (3.5) | 3.7 (4.3) | | 1.4 (2.6) |
| Mean ESR (mm/h) | | 31.4 (21.6) | 36.8 (21.3) | | 27.1 (23.0) |
| Elevated inflammatory marker [3] | | 31 (69%) | 18 (90%) | | 19 (58%) |
| Mean albumin (g/dL) | | 4.1 (0.7) | 4.1 (0.5) | | 4.3 (0.6) |
| Hypoalbuminaemia (<3.5 g/dL) | | 6 (13%) | 2 (10%) | | 2 (6%) |
| Mean hemoglobin (g/dL) | | 11.9 (1.8) | 11.7 (1.6) | | 11.4 (2.6) |
| Anemia (<10 g/dL) | | 6 (13%) | 3 (15%) | | 8 (24%) |
| Median fecal calprotectin (mg/L) (IQR) | | 954 (166 to 3041) | 2068 (825 to 7362) | | 1141 (491 to 2972) |
| **Treatment** | | | | | |
| Newly diagnosed | | 30 (67%) | 20 (100%) | | 16 (48%) |
| EEN therapy duration in days | | | 52 (19) | | n.a. |
| Previous therapies | EEN | 11 (%) | - | | n.a. |

| | | | | | |
|---|---|---|---|---|---|
| | 5-ASA | 2 (4%) | - | | 4 (12%) |
| | Steroids | 8 (18%) | - | | 9 (27%) |
| | IM | 7 (16%) | - | | 1 (3%) |
| | Previous biologics | 3 (7%) | - | | 1 (3%) |
| Ongoing therapies at time of study recruitment | 5-ASA | 4 (9%) | - | | 13 (39%) |
| | Steroids | 1 (2%) | - | | 4 (12%) |
| | IM | 4 (9%) | - | | 3 (9%) |
| | Anti-TNF | 5 (11%) | - | | 3 (9%) |

***Table S1: Patient characterization***

*Data are N (%), mean (SD), or median (IQR 25th to 75th percentile). 5-ASA, 5-aminosalicylic acids; BMI, body-mass index; CRP, C-reactive protein; EEN, exclusive enteral nutrition; ESR, erythrocyte sedimentation rate; IBD, inflammatory bowel disease; IM, immunomodulater (e.g., azathioprine, methotrexate); PUCAI, Pediatric Ulcerative Colitis Activity Index; TNF, tumor necrosis factor; wPCDAI, weighted Pediatric Crohn's Disease Activity Index. [1] As per Paris classification. [2] Five patients had joint manifestations, two had eye manifestations, eight had skin manifestations, one had autoimmune sclerosing cholangitis and one had autoimmune hepatitis. [3] CRP ≥ 0.5 mg/L or ESR ≥ 25 mm/h.*

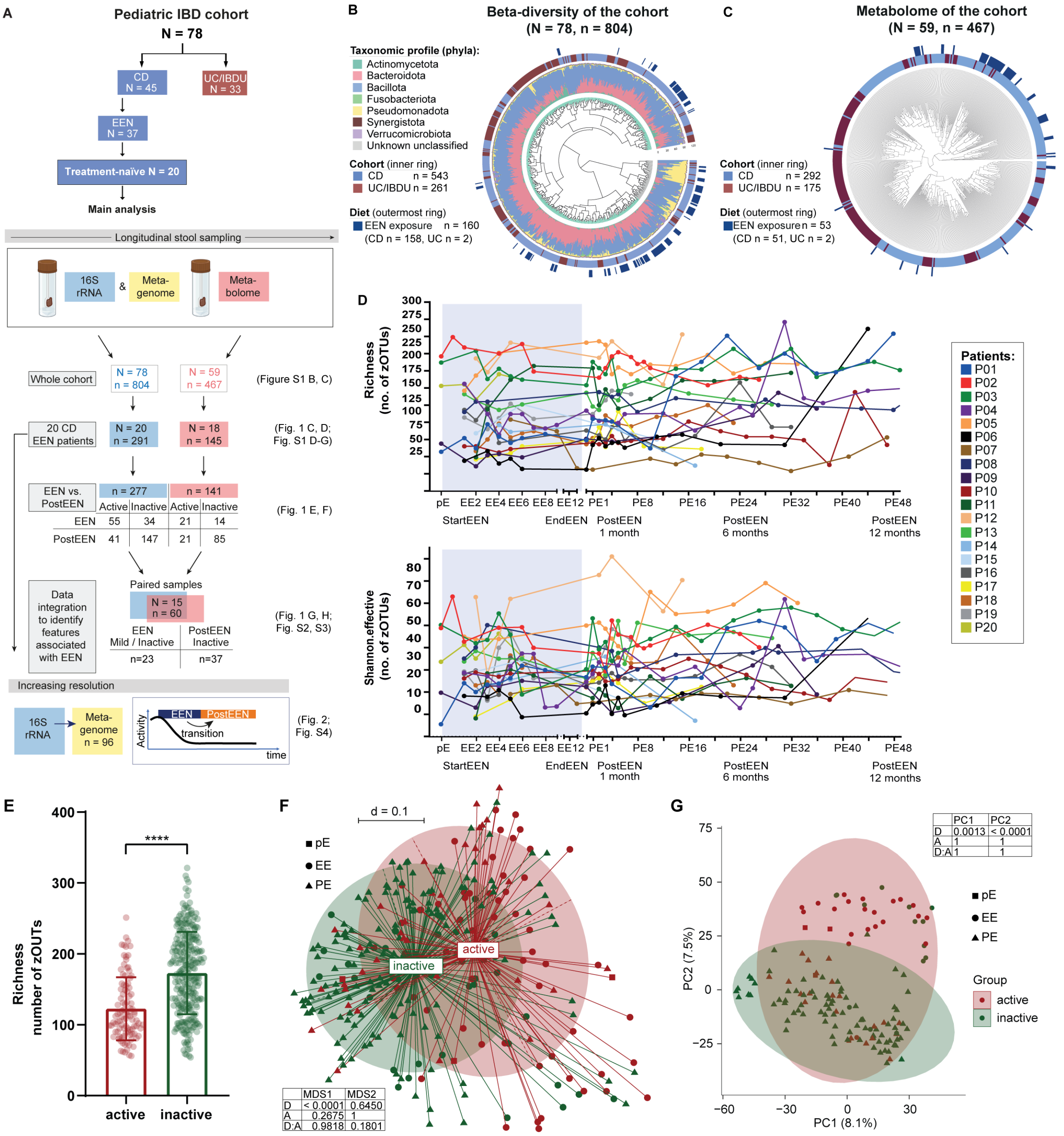


***Figure S1: Overview of the pediatric IBD cohort, sampling and analysis and Alpha-diversity in CD-EEN***

***A** Overview of the pediatric IBD cohort (33 UC/IBDU, 45 CD patients), focusing on 20 newly diagnosed and treatment-naïve CD patients receiving Exclusive Enteral Nutrition (EEN) for the first time. We longitudinally collected fecal samples for 16S rRNA amplicon and metagenomic sequencing as well as metabolome analysis. Overview of analysis (1-4) is shown with quantity of patients (N) and samples (n). **B** Beta-diversity analysis of the fecal microbiota in IBD patients. The circular dendrogram shows differences in beta-diversity from the longitudinal microbial profiling based on generalized UniFrac distances between 804 samples of 78 patients of the cohort. Taxonomic composition of fecal samples at the phylum level is shown as color-coded stacked bar blots around the dendrogram. Bars in the first outer ring of the figure show disease phenotypes (CD, UC/IBDU) and EEN treatment in the second outer ring. **C** Complete clustering of untargeted metabolomic samples with Euclidean distance for 467 longitudinal fecal samples from 59 IBD patients, visualized in a circular tree. **D** Alpha-diversity plotted as richness (number of zOTUs) and Shannon effective (number of zOTUs) for all samples from the 20 newly diagnosed pCD patients undergoing EEN therapy. **E** Richness (number of zOTUs) in samples from patients in active or inactive disease based on pga (physician global assessment, active = severe, moderate, mild, remission = remission).*

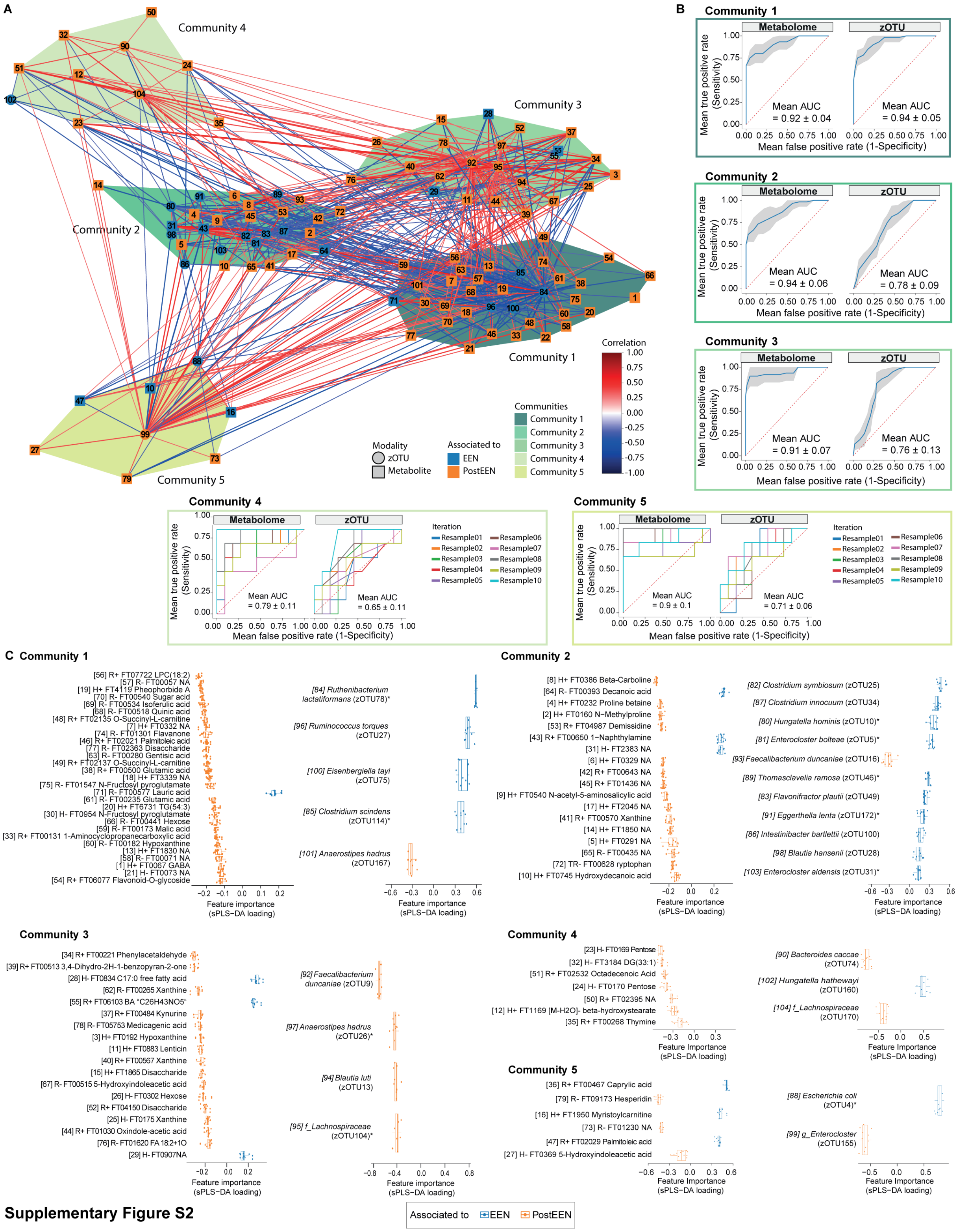


**Supplementary Figure S2**

***Figure S2: Data integration and identification of features of EEN:***

***A** Correlation network built from selected features based on sPLS-DA-based data integration. Five communities were identified (color coded as indicated). Indicated are zOTUs (circles) and metabolites (squares), their associations to EEN (blue) or PostEEN (orange), and correlation values (lines between features, blue to red). **B** ROC curves for metabolome and zOTU features, respectively, for Community 1, 2, and 3, grey area ±1 SD. All curves shown were computed on held-out test samples not seen during model fitting. **C** Most important features (i.e., metabolites and zOTUs) for each community with their feature importance for PostEEN (<0, orange) and EEN (>0, blue). One dot represents the loading of the respective feature for a single iteration of the cross-validation* *procedure (i.e. each iteration is trained on a (stratified) random subset of all data points). *BONCAT responders Fig.2.*

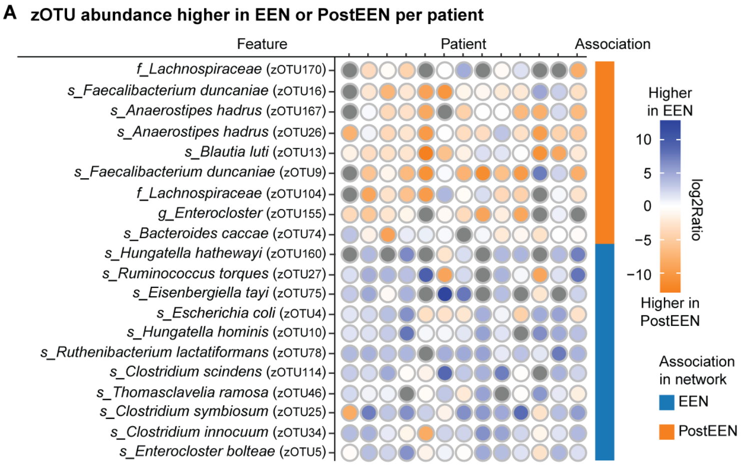


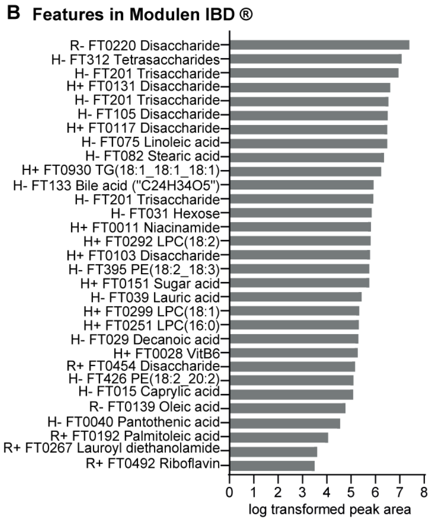


***Figure S3: patient-specific expression levels of EEN and PostEEN zOTU signatures***

***A** Dotplot of patient-specific expression levels of EEN and PostEEN zOTU signatures (N = 13; all of which EEN and PostEEN samples were available) showing individual log fold-changes between expression levels of EEN and PostEEN samples. The mean of expression was used when more than one sample was available per patient and condition, i.e. EEN or PostEEN. Blue, higher expression in EEN samples; orange, higher expression levels in PostEEN samples; white, balanced between conditions; grey, zOTU was not present in the EEN and PostEEN sample. zOTUs identified with SILVA in the network analyses were re-checked using 16S-based ID tool of EzBioCloud[56] using the database update 2023.08.23, reflecting all taxa published and accepted by IJSEM until April 2023. **B** Identified features of Modulen IBD® from untargeted metabolomic analyses.*

***Table S2: zOTU identification with EZBioCloud***

*zOTUs identified with SILVA in the network analyses were re-checked using the 16S-based ID tool of EzBioCloud[56] using the database update 2023.08.23, reflecting all taxa published and accepted by IJSEM until April 2023. d Identified features of Modulen IBD® from untargeted metabolomic analyses. Previously members of Lachnoclostridium are highlighted in bold. *zOUTs identified as BONCAT responders Fig.2, [+] zOUTs identified as differential abundant in mouse experiments Fig. 5D-F.*

| **Associated to** | **zOUT** | **EZBioCloud** | | | **genus (SILVA database (Quast et al., 2013))** |
|---|---|---|---|---|---|
| | | **species** | **family** | **genus** | |
| EEN | 78*[+] | *Ruthenibacterium lactatiformans* | *Oscillospiraceae* | *Ruthenibacterium* | *UBA1819* |
| **EEN** | **114*** | ***Clostridium scindens*** | ***Lachnospiraceae*** | ***Sporofaciens*** | ***Lachnoclostridium*** |
| EEN | 27[+] | *Ruminococcus torques* | *Lachnospiraceae* | *Mediterraneibacter* | *Ruminococcus torques group* |
| EEN | 75 | *Eisenbergiella tayi* | *Lachnospiraceae* | *Eisenbergiella* | *Eisenbergiella* |
| EEN | 10* | *Hungatella hominis* | *Lachnospiraceae* | *Hungatella* | *f_Lachnospiraceae* |
| **EEN** | **5*[+]** | ***Enterocloster bolteae*** | ***Lachnospiraceae*** | ***Enterocloster*** | ***Lachnoclostridium*** |
| **EEN** | **25[+]** | ***Clostridium symbiosum*** | ***Lachnospiraceae*** | ***Clostridium_g35*** | ***Lachnoclostridium*** |
| EEN | 49[+] | *Flavonifractor plautii* | *Oscillospiraceae* | *Pseudoflavonifractor* | *Flavonifractor* |
| EEN | 100 | *Intestinibacter bartlettii* | *Peptostreptococcaceae* | *Intestinibacter* | *Intestinibacter* |
| EEN | 34[+] | *Clostridium innocuum* | *Erysipelotrichaceae* | *Clostridium_g36* | *Clostridium innocuum group* |
| EEN | 46*[+] | *Thomasclavelia ramosa* | *Coprobacillaceae* | *Thomasclavelia* | *Erysipelatoclostridium* |
| EEN | 172* | *Eggerthella lenta* | *Eggerthellaceae* | *Eggerthella* | *Eggerthella* |
| EEN | 28 | *Blautia hansenii* | *Lachnospiraceae* | *Blautia* | *Blautia* |
| **EEN** | **31*** | ***Enterocloster aldensis*** | ***Lachnospiraceae*** | ***Enterocloster*** | ***Lachnoclostridium*** |
| EEN | 160 | *Hungatella hathewayi* | *Lachnospiraceae* | *Hungatella* | *Hungatella* |
| EEN | 4* | *Escherichia coli* | *Enterobacteriaceae* | *Escherichia* | *Escherichia-Shigella* |
| PostEEN | 167 | *Anaerostipes hadrus* | *Lachnospiraceae* | *Anaerostipes* | *Anaerostipes* |
| PostEEN | 16 | *Faecalibacterium duncaniae* | *Oscillospiraceae* | *Faecalibacterium* | *Faecalibacterium* |
| PostEEN | 9 | *Faecalibacterium duncaniae* | *Oscillospiraceae* | *Faecalibacterium* | *Faecalibacterium* |
| PostEEN | 13[+] | *Blautia luti* | *Lachnospiraceae* | *Blautia* | *Blautia* |
| PostEEN | 104* | *QUHQ_s* | *Lachnospiraceae* | *QUHQ_g* | *f_Lachnospiraceae* |
| PostEEN | 26* | *Anaerostipes hadrus* | *Lachnospiraceae* | *Anaerostipes* | *Anaerostipes* |
| PostEEN | 74 | *Bacteroides caccae* | *Bacteroidaceae* | *Bacteroides* | *Bacteroides* |
| PostEEN | 170 | *EU459701_s* | *Lachnospiraceae* | *QTXI_g* | *Lachnospiraceae NK4A136 group* |
| **PostEEN** | **155** | ***PAC001295_s*** | ***Lachnospiraceae*** | ***Enterocloster*** | ***Lachnoclostridium*** |

***Table S3: Overview of available samples in glycerol from MP1-MP3 with respective patient clinical data and information on fecal sample and transfer in model systems.***

<table>
<tr><td></td><td>Patient</td><td colspan="6">MP1</td><td colspan="5">MP2</td><td colspan="3">MP3</td></tr>
<tr><td>Sampling timepoint</td><td>Sample along therapy</td><td>PreEEN</td><td colspan="3">EEN</td><td colspan="2">PostEEN</td><td>PreEEN</td><td colspan="2">EEN</td><td>PostEEN (4 weeks before relapse)</td><td>PostEEN (5 weeks after relapse)</td><td>PreEEN</td><td>EEN</td><td>PostEEN</td></tr>
<tr><td></td><td>Sampling timepoint [weeks]</td><td>pE</td><td colspan="3">EE4</td><td colspan="2">PE10</td><td>pE</td><td colspan="2">EE6</td><td>PE4</td><td>PE14</td><td>pE</td><td>EE6</td><td>PE36</td></tr>
<tr><td>Clinical activity</td><td>pga</td><td>moderate</td><td colspan="3">remission</td><td colspan="2">remission</td><td>severe</td><td colspan="2">remission</td><td>remission</td><td>mild</td><td>moderate</td><td>remission</td><td>remission</td></tr>
<tr><td></td><td>Calprotectin [mg/L]</td><td>1684</td><td>3121 (EE3)</td><td>NA</td><td>103 (EE5)</td><td colspan="2">26</td><td>NA</td><td>1994 (EE3)</td><td>NA</td><td>400</td><td>270</td><td>> 8000</td><td>2828</td><td>33</td></tr>
<tr><td></td><td>wPCDAI</td><td>45</td><td>0 (EE2)</td><td colspan="2">NA</td><td>0 (PE5)</td><td>NA</td><td>70</td><td>10 (EE2)</td><td>NA</td><td>0</td><td>22.5</td><td>52.5</td><td>7.5</td><td>10</td></tr>
<tr><td>Drug exposure</td><td>Co-medication</td><td>-</td><td colspan="3">Anti-TNF</td><td colspan="2">Anti-TNF</td><td>-</td><td colspan="2">MTX</td><td>MTX</td><td>Anti-TNF</td><td>-</td><td>MTX</td><td>Anti-TNF</td></tr>
<tr><td></td><td>Time under co-medication [days]</td><td>-</td><td colspan="3">9</td><td colspan="2">98</td><td>-</td><td colspan="2">2</td><td>41</td><td>30</td><td>-</td><td>24</td><td>96</td></tr>
<tr><td>Sample quality</td><td>Bacterial biomass [reads per gram feces]</td><td>81149</td><td colspan="3">13664</td><td colspan="2">38813</td><td>1846</td><td colspan="2">11087</td><td>34184</td><td>42872</td><td>33081</td><td>63210</td><td>101750</td></tr>
<tr><td>Transfer</td><td>Mouse FMT 1 Il10$^{-/-}$ phenotype (Colon mean HS)</td><td>Inflamed (5.3)</td><td colspan="3">Inflamed (4.8)</td><td colspan="2">Inflamed (3.7)</td><td>Not inflamed (1.7)</td><td colspan="2">Not inflamed (1.8)</td><td>Not inflamed (2.5)</td><td>Inflamed (4.3)</td><td>Inflamed (8.5)</td><td>Inflamed (7.3)</td><td>Inflamed (5.3)</td></tr>
<tr><td colspan="2">Selected for subsequent experiments</td><td>yes</td><td colspan="3">no</td><td colspan="2">no</td><td>no</td><td colspan="2">yes</td><td>no</td><td>yes</td><td>no</td><td>yes</td><td>no</td></tr>
</table>

*Information visualized also in Fig. 9, 11 and Supplementary Fig 3. Sampling timepoint [weeks]: pE = PreEEN before start of EEN therapy; EE4 = sample collected in fourth week of EEN therapy; PE10 = sample collected ten weeks after end of EEN (PostEEN). NA = Data not available; - = no co-medication; wPCDAI = weighted paediatric Crohn's disease activity index; pga = physician global assessment. HS = Histopathology score. If data is not available from the selected timepoint, a reference timepoint before or after, where the data is available, is given.*

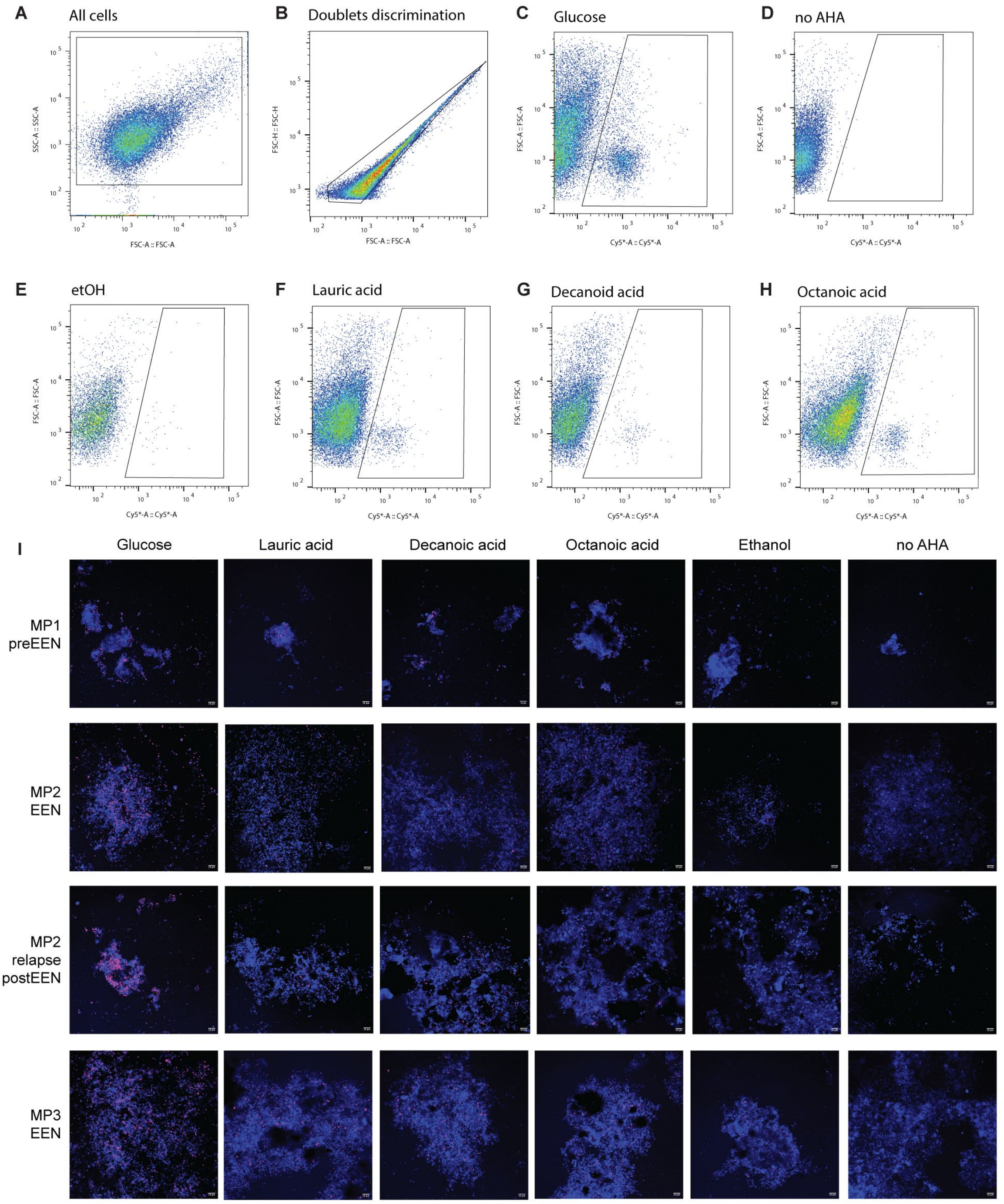


***Figure S4: BONCAT-FACS gating strategy and microscopy pictures depicting translationally active bacterial cells:***

*Bacteria were sorted with FACS Melody cell sorter. The sorting gate was utilized to isolate the active fraction after incubation with different medium chain fatty acids (MCFAs).* ***A*** *Bacteria were visualized using the forward scatter (FSC) and side scatter (SSC) and pre-gated.* ***B*** *Doublet discrimination was conducted to exclude doublets from analysis, preventing false positives in the sorted fraction.* ***C*** *positive control with glucose was included in every experiment to verify the accuracy of the click reaction and active cell labeling.* ***D*** *Stool sample incubated without AHA was used as negative control and to select the gate position.* ***E*** *Ethanol was used as negative control for the MCFAs, (****F****) Cy5-labeled cells in the presence of* ***F*** *lauric acid,* ***G*** *decanoic acid, and* ***H*** *octanoic acid.* ***I*** *Representative confocal microscopy images of stool samples from the patients MP1 preEEN, MP2 EEN, MP2 relapse and MP3 EEN stimulated with MCFAs (lauric acid, decanoic acid and octanoic acid) under anaerobic* *conditions. Active bacterial cells are shown in pink (BONCAT positive signal), while all cells are shown in blue (DAPI). Due to MCFAs solubility in ethanol, ethanol was used as negative control. Additionally, stool samples were incubated without AHA to ensure that there was no background noise in the Cy5 channel. Scale bar is 10μm.*

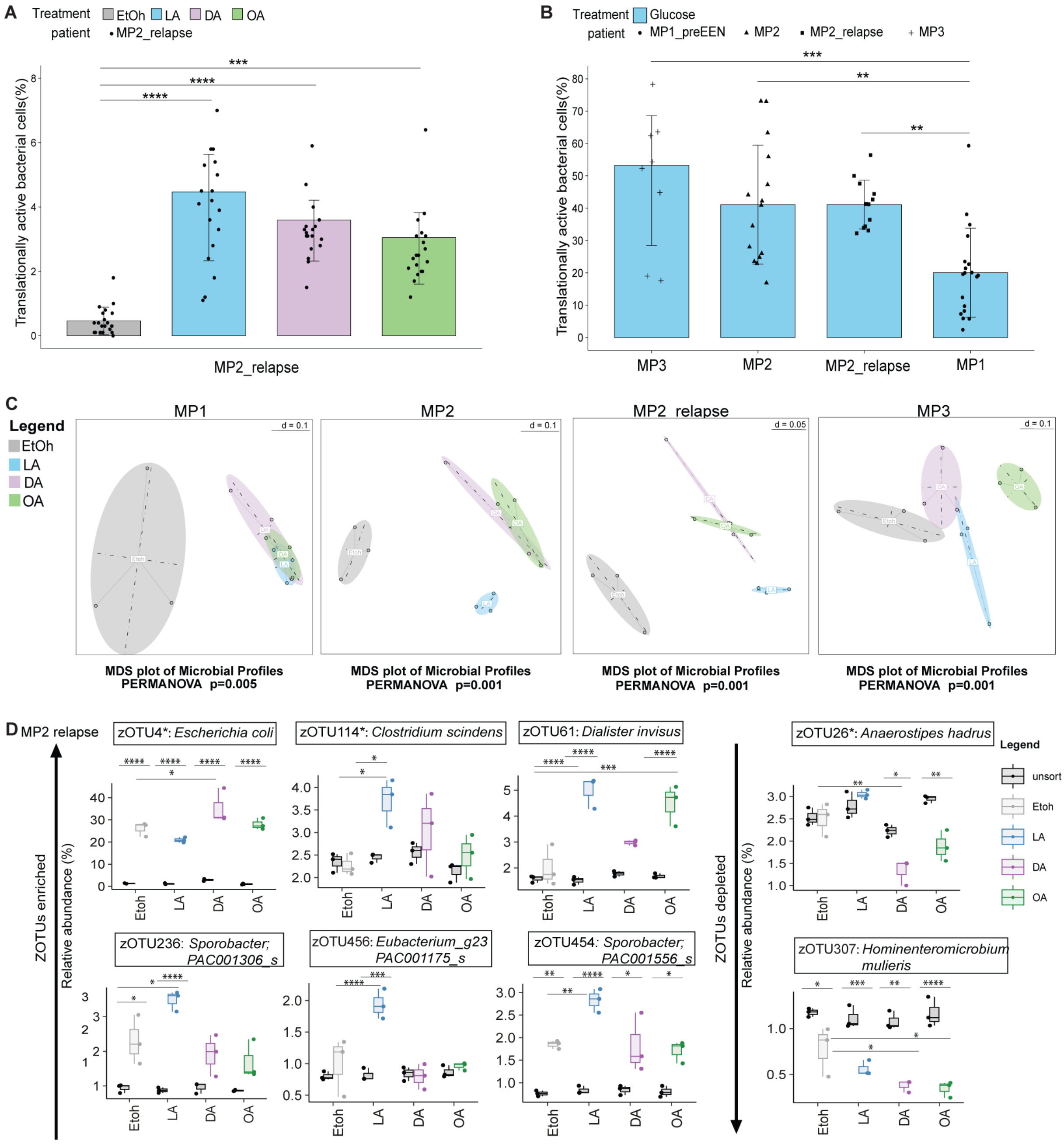


***Figure S5: BONCAT results for MP2_relapse sample with beta-diversity MDS plots of the sorted fraction:***
***A** Translationally active bacterial cells in MP2_relapse patient (ethanol, n=20; LA, n=19; DA, n=19 and OA n=20).* ***B** Translationally active bacterial cells in glucose. Glucose was used as positive control in all experiment and measured in all patients (MP1, n=19; MP2, n=15; MP2_relapse, n=12 and MP3, n=10). P values were calculated by Kruskal-Wallis test and Dunn test for multiple comparisons. *p < 0.05, **p < 0.01, ***p < 0.001, ****p < 0.0001.* ***C** Beta-diversity MDS plots shows clustering of samples based on different amendments (MCFAs and ethanol) in the sorted fraction (perMANOVA).* ***D** Relative abundances of significant enriched and depleted zOTUs in MP2 relapse. *zOUTs identified in Network analyses Fig 1G, S2. P values were calculated by ANOVA and Tukey's test for multiple comparisons. *p < 0.05, **p < 0.01, ***p < 0.001, ****p < 0.0001.*

***Table S4. List of significant zOTUs enriched in the sorted fraction after exposure to MCFAs.***

| Patient | Enriched zOTUs | Identification | Similarity (%) | MCFAs |
|---|---|---|---|---|
| **MP1** | zOTU 4 | *Escherichia coli* | 100 | LA, DA, OA |
| | zOTU 104 | *Lachnospiraceae QUHQ_s*<br>*Enterocloster bolteae* | 100<br>95 | OA |
| **MP2** | zOTU 5 | *Enterocloster bolteae* | 100 | LA, DA, OA |
| | zOTU 31 | *Enterocloster aldensis* | 99.5 | LA, DA, OA |
| | zOTU 930 | *Enterocloster bolteae* | 99.5 | OA |
| | zOTU 172 | *Eggerthella lenta* | 100 | LA |
| **MP2_relapse** | zOTU 4 | *Escherichia coli* | 100 | DA |
| | zOTU 114 | *Clostridium scindens* | 100 | LA |
| | zOTU 61 | *Dialister invisus* | 100 | LA,OA |
| | zOTU 236 | *Sporobacter;* PAC001306_s | 100 | LA |
| | zOTU 456 | *Eubacterium_g23*; PAC001175_s<br>*Eubacterium coprostanoligenes* | 99.5<br>95.7 | LA |
| | zOTU 454 | *Sporobacter*; PAC001556_s | 99 | LA |
| **MP3** | zOTU 4 | *Escherichia coli* | 100 | OA |

*zOTUs were identified using the 16S-based ID tool of EzBioCloud.*

***Table S5. List of significant zOTUs depleted in the sorted fraction after exposure to MCFAs.***

| **Patient** | **Enriched zOTUs** | **Identification** | **Similarity (%)** | **MCFAs** |
|---|---|---|---|---|
| **MP2** | zOTU 114 | *Clostridium scindens* | 100 | DA |
| | zOTU 46 | *Thomasclavelia ramosa* | 100 | OA |
| | zOTU 10 | *Hungatella hominis* | 100 | OA |
| | zOTU 78 | *Ruthenibacterium lactatiformans* | 100 | OA |
| | zOTU 443 | *Peptostreptococcus anaerobius* | 100 | DA |
| | zOTU 1 | *Phocaeicola vulgatus* | 99.5 | OA |
| **MP2_relapse** | zOTU 26 | *Anaerostipes hadrus* | 100 | DA |
| | zOTU 307 | *Hominenteromicrobium mulieris* | 100 | DA, OA |

*zOTUs were identified using the 16S-based ID tool of EzBioCloud.*

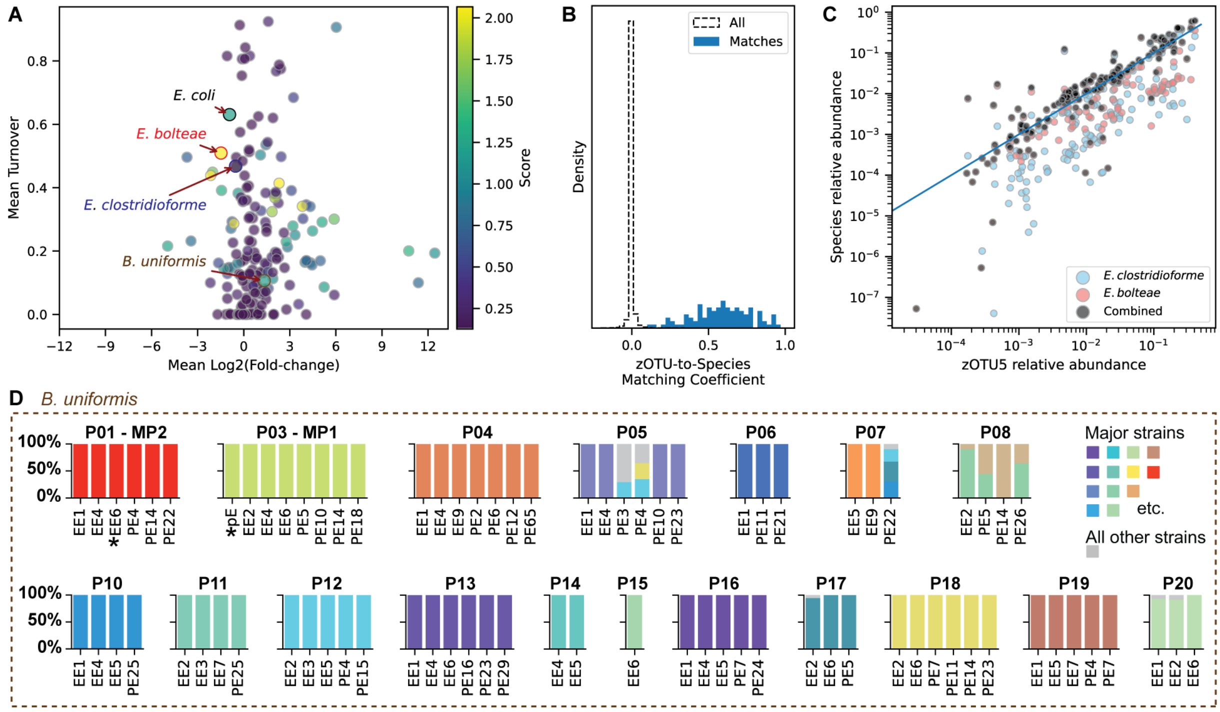

***Figure S6: Species and strain turnover during the transition off of EEN***

***A** Characterization of species based on fold-change in relative abundance between EEN and PostEEN samples and average strain-level turnover, defined as the mean Bray-Curtis dissimilarity between within-subject sample pairs. Particularly interesting species were identified based on the product of these two statistics, the number of within-subject sample pairs, and the mean relative abundance (**Supplementary Table 6**). Points are colored based on this score. Highlighted species (colored circles) presented in detail in **Fig. 3D-F**, and **D**: Composition over time of strains within* E. bolteae *(red),* E. clostridioformis *(blue),* E. coli *(black) and **S6D*** Bacteroides uniformis *(brown) in the CD-EEN patients. **B** The distribution of correlations between zOTUs and species. In this way, 89 metagenomic species were matched to 95 zOTUs, accounting for a median of 74% of relative abundance per sample. **C** Correlation between the relative abundance of zOTU5 and the individual and combined relative abundances of the species* Enterocloster bolteae *and* Enterocloster clostridioformis *determined by quantification with shotgun metagenomes. The blue diagonal represents a 1:1 relationship in relative abundance. **D** Composition over time of strains within* Bacteroides uniformis *abundant in the CD-EEN patients. Strain genotypes and fractions were estimated from shotgun metagenomic data. Fractions of abundant strains (colored bars) are shown across samples from individual subjects (P01, P02…), before EEN (“pE” labels on x-axis), during (“EE”) and after (“PE”). * Samples used as donor samples in other experiments. Strain fractions for the species sum to 1; minor strains are summed together and shown in grey*

***Table S6: Matching of metagenomic and 16S rRNA sequencing results***

*See excel file. Identified candidate matches where species (based on ubiquitous marker genes) and zOTU relative abundance profiles were associated across samples. Here, 89 metagenomic species were matched to 95 zOTUs, accounting for a median of 74% of relative abundance per sample.*

***Table S7: Identification of 46 species with extensive strain turnover, large changes in relative abundance at the end of EEN, high mean relative abundance, and high prevalence***

*See excel file. Indicator_score was calculated as = |species_log2_fold_change|* species_overall_mean_relative_abundance* num_intrasubject_sample_pairs * mean_pairwise_braycurtis_ dissimilarity. P-values for species enrichment ("species_log2_fold_change") were calculated using a Wilcoxon signed-rank test. The effect of the transition on strain turnover ("transition_turnover_effect") is the difference between the mean time and subject-adjusted pairwise Bray-Curtis dissimilarities for pairs of samples spanning the transition compared to pairs during each of the EEN or PostEEN time periods. The P-value for this effect is estimated using a permutation test (n=999).*

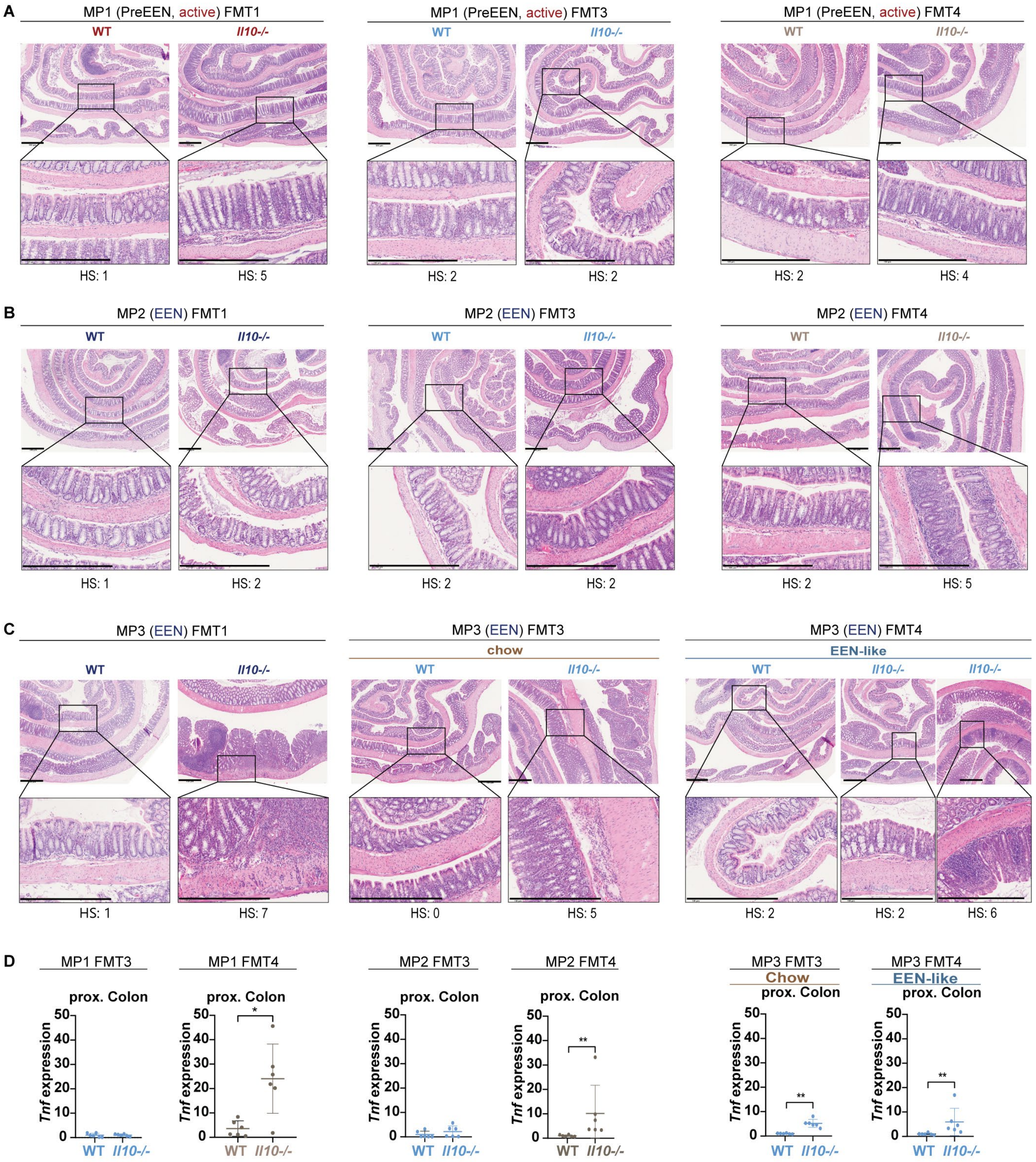


***Figure S7: Histopathology of mouse transfers:***

***A-C:*** *Mouse transfer experiments: Representative (mean of group) H&E stained sections of colonic Swiss roles and corresponding higher magnifications for Il10-/- mice and respective controls from FMT1, 3 and 4 (scale bars = 500 µM, HS = histopathology score).* ***A****: MP1.* ***B****: MP2.* ***C****: MP3.* ***D*** Tnf *gene expression levels (fold of control) in proximal colon. Data are represented by mean ± SD of six biological replicates. P values were calculated by Mann-Whitney test. *p < 0.05, **p < 0.01, ***p < 0.001, ****p < 0.0001.*

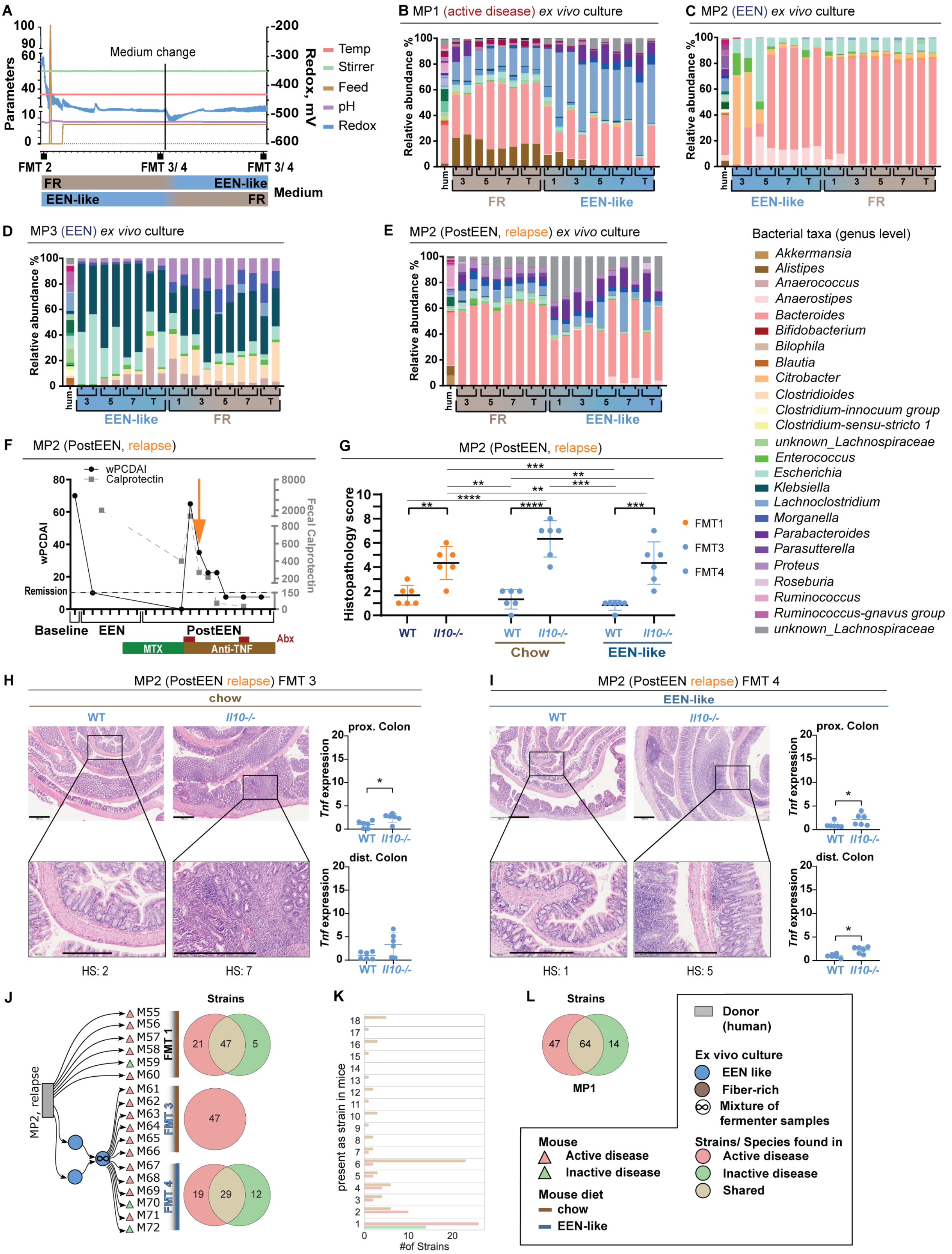

***Figure S8:* Ex vivo *gut chemostat and MP2 relapse transfers***

***A** Parameters of* ex vivo *gut model over time with inoculation (FMT 2) and transfer time points into mice (FMT 3 and 4). Depending on the sample's origin, the start media is chosen to match the diet of the sample at time of collection (PreEEN on FR media (brown), EEN on EEN-like media (blue)) with a switch to the respective other media in between. Parameters are recorded over the full course of the experiment and plotted as seen: feed, pH, temperature, stir speed and redox potential.* ***B-E** 16S rRNA gene sequencing of* ex vivo *continuous culture of human (hum) fecal sample using different media performed in one experiment with transfer (timepoint, T) into the mouse* *model at the end of each media exposure (equals FMT 3 and 4), FR = brown, EEN-like = blue.* ***B*** *MP1, PreEEN active disease.* ***C*** *MP2, EEN.* ***D*** *MP3, EEN.* ***E*** *MP2, relapse.* ***F*** *Selected MP2, relapse sample for transfer: patients' disease state with wPCDAI and fecal calprotectin, therapy response over time, maintenance therapy (MTX, Anti-TNF) and sampling time point of transferred fecal sample (orange arrow).* ***G*** *Mouse histopathology scoring of colon Swiss roles of FMT 1, 3 and 4 from MP2, relapse.* ***H, I*** *Mouse transfer experiments from MP2 relapse: Representative (mean of group) H&E stained sections of colonic Swiss roles and corresponding higher magnifications for $Il10^{-/-}$ mice and respective controls (scale bars = 500 µM, HS = histopathology score) with* Tnf *gene expression levels (fold of control) in proximal and distal Colon.* ***H*** *MP2 relapse FMT 3, chow diet.* ***I*** *MP2 relapse FMT 4, EEN-like diet. J Transfer overview of MP2 relapse donor sample (grey) from FMT 1 (black), FMT 3 (EEN-conditioned, blue) and FMT 4 (FR-conditioned, brown) into $Il10^{-/-}$ mice. Mouse inflammation endpoints are shown (triangles, red = active, green = inactive). Venn diagrams tally strains detected in inflamed (red), non-inflamed (green) $Il10^{-/-}$ mice or found in both conditions (yellow).* ***K*** *Histogram shows presence of strains in all 18 Il10-/- mice from FMT1, 3 and 4 for each MP2, relapse sample.* ***L*** *Venn diagrams combining strains across FMT1, 3, and 4 for MP2 relapse sample. Data are represented by (**G, H, I**) mean ± SD of six biological replicates. P values were calculated by Mann-Whitney test (**H, I**) or two-way ANOVA with Tukey multiple pairwise-comparisons test (**G**). *p < 0.05, **p < 0.01, ***p < 0.001, ****p < 0.0001.*

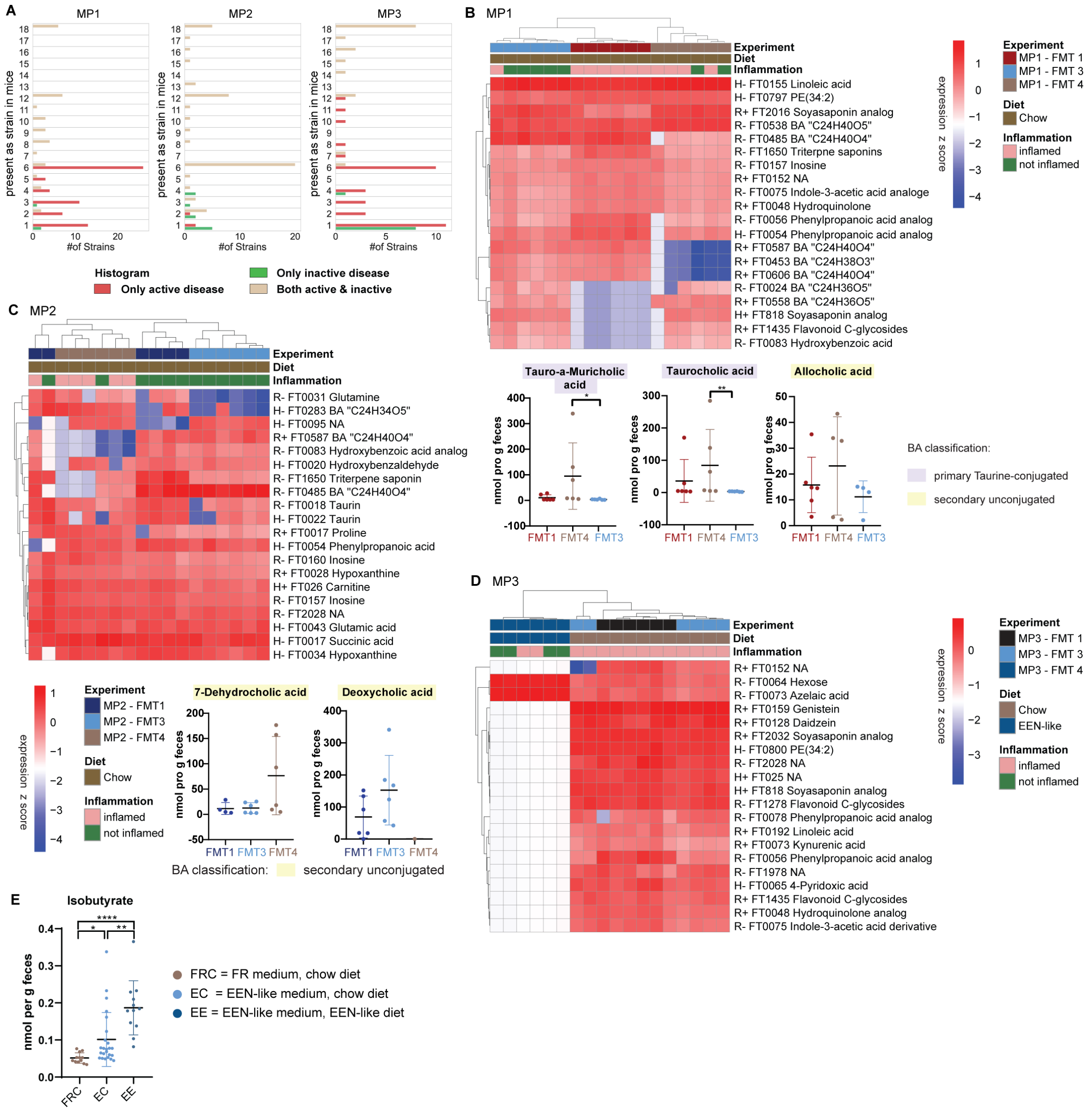


***Figure S9: Functional difference in Il10$^{-/-}$ mice***

***A** Histogram shows presence of strains in all 18 Il10-/- mice from FMT1, 3 and 4 for each MP. **B-D** Heatmap of differential abundant metabolites based on inflammation and FMT. **B** MP1 with targeted BA (bile acids) validation. **C** MP2 with targeted BA (bile acids) validation. **D** MP3. **E** Targeted results of Isobutyrate concentration in Il10-/- mice from FMT 3 and 4 for MP1 – 3. Mice colonized with: FRC = FR medium conditioned microbiota, mice under chow diet, EC = EEN-like medium conditioned microbiota, mice under chow diet, EE = EEN-like medium conditioned microbiota, mice under EEN-like diet. BA data are represented by mean ± SD of six biological replicates. NA values were not counted as 0. P-values were calculated by ANOVA followed by Dunn's multiple comparison test. *p < 0.05, **p < 0.01, ***p < 0.001, ****p < 0.0001. Histopathology score > 3 was considered as inflamed phenotype.*

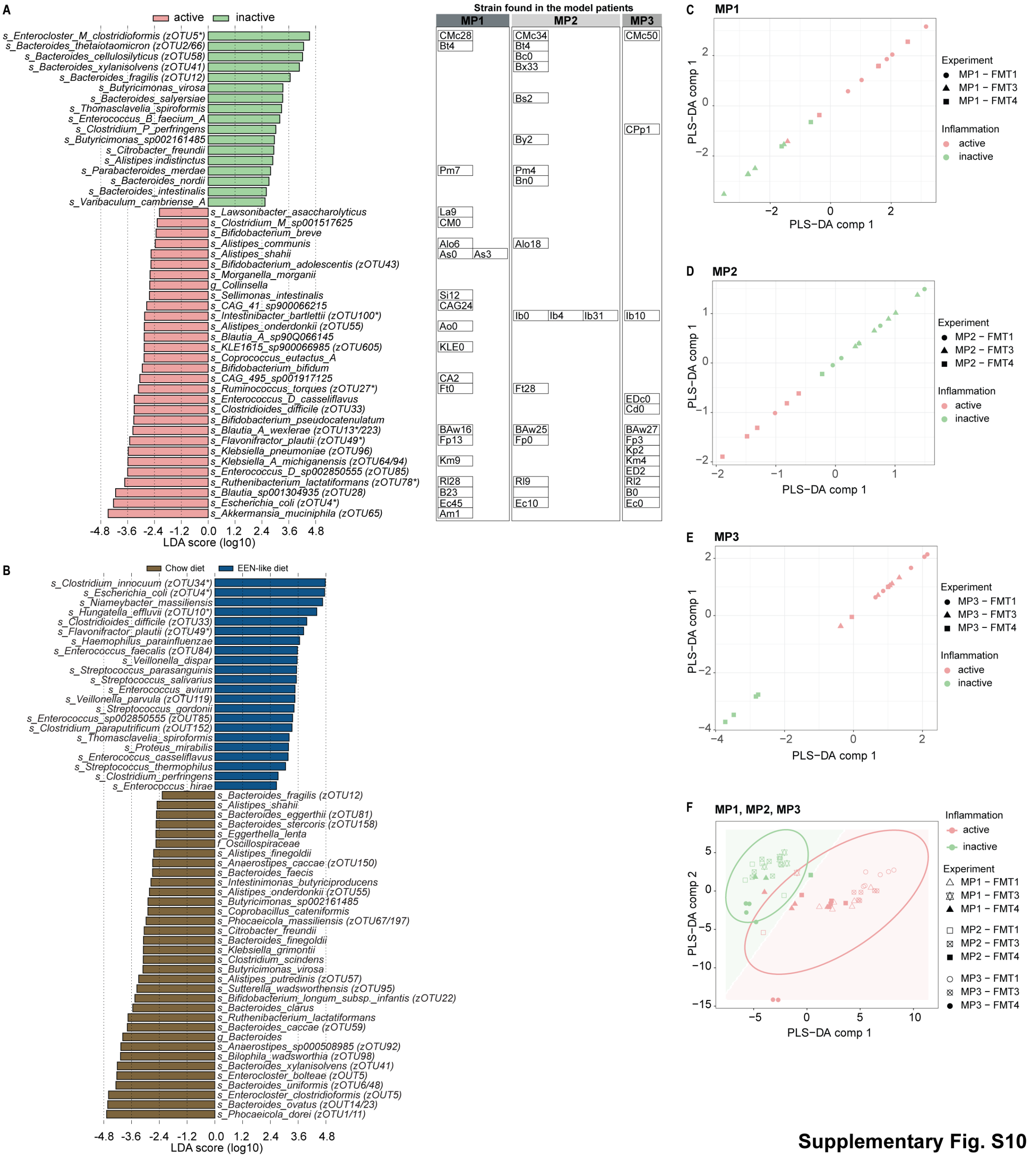


**Supplementary Fig. S10**

***Figure S10: Strains associated with active or inactive disease in Il10$^{-/-}$ mice***:

***A*** *LEfSe (Linear discriminant analysis Effect Size) analysis of differentially abundant bacteria at species level in all recipient inflamed (active, red) or non-inflamed (inactive, green) Il10$^{-/-}$ mice from all FMTs performed with abundance of strains in each MP. Different strains are labeled with different numbers.* ***B*** *LEfSe analysis of differentially abundant bacteria at species level based on metagenomic data in mouse experiments from MP1- MP3 (including relapse) based on mouse diet (chow – brown, EEN-like – blue).* ***C-E*** *Analyses of untargeted metabolomics with representative sPLS-DA plots comparing inflammation in Il10$^{-/-}$ mice (inflamed = red; not inflamed = green) from FMT 1, 3 and 4. Axes indicate the sPLS-DA component separating the groups.* ***C*** *MP1.* ***D*** *MP2.* ***E*** *MP3.* ***F*** *Visualization of inflamed and non-inflamed mice on the two identified sPLS-DA components with samples from FMT1, 3 and 4 from transfers from MP1-3. Strain abundance cut-off 20%, species abundance cut-off $10^{-5}$, prevalence ≥ 1. Histopathology score > 3 was considered as inflamed phenotype = active. *zOTUs found in Network analyses Fig 1G, S2.*

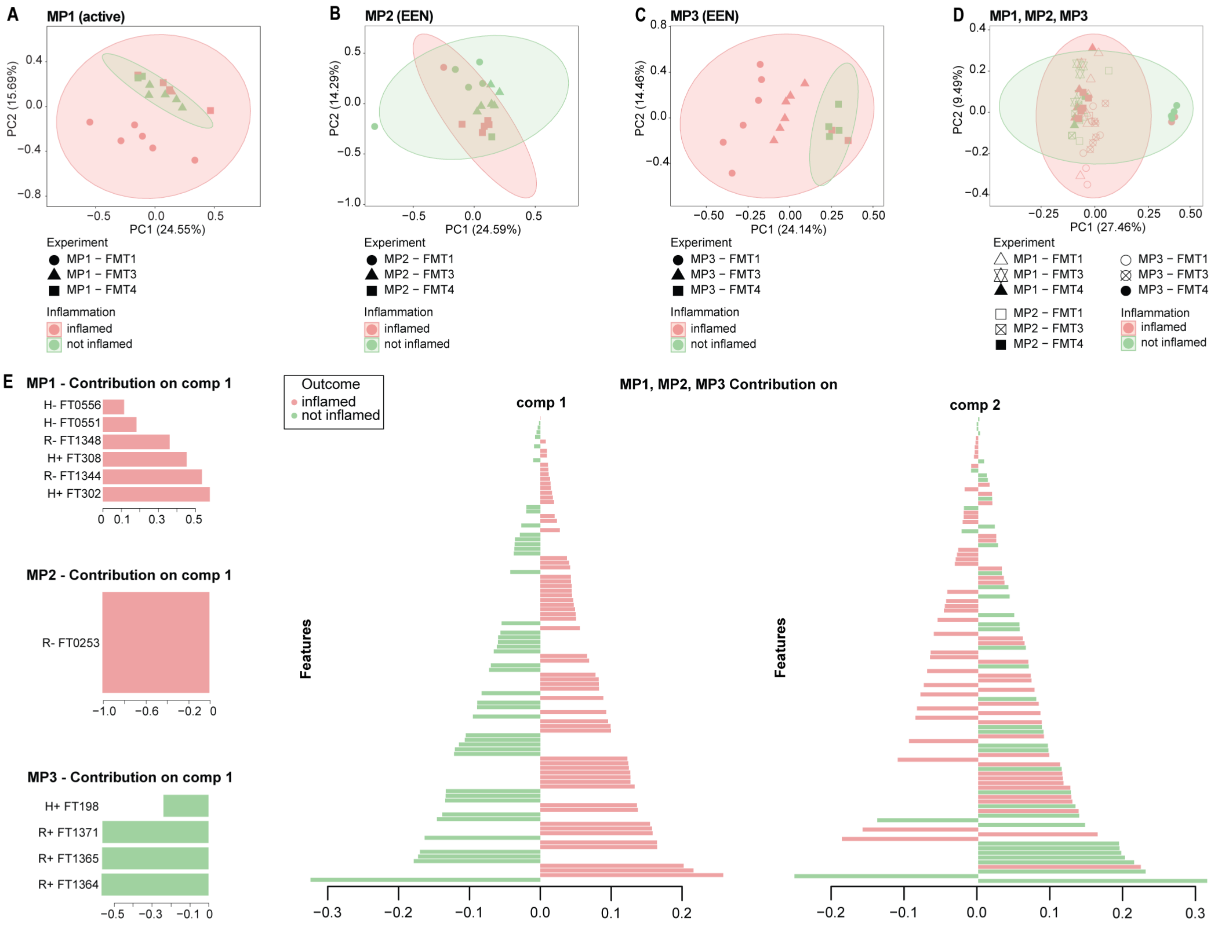


***Figure S11: Untargeted metabolomics in all transfer experiments***

***A-D** Representative PCA plots comparing inflammation in Il10$^{-/-}$ mice (inflamed = red; not inflamed = green) from FMT 1, 3 and 4. Axes indicate principal components (PC), with PC1 representing the most variation (%) and PC2 representing the second most variation (%). **A** MP1. **B** MP2. **C** MP3. **D** MP1, MP2 and MP3. **E** Raw values of top VIP features comparing both conditions. (Histopathology score > 3 was considered as inflamed phenotype).*

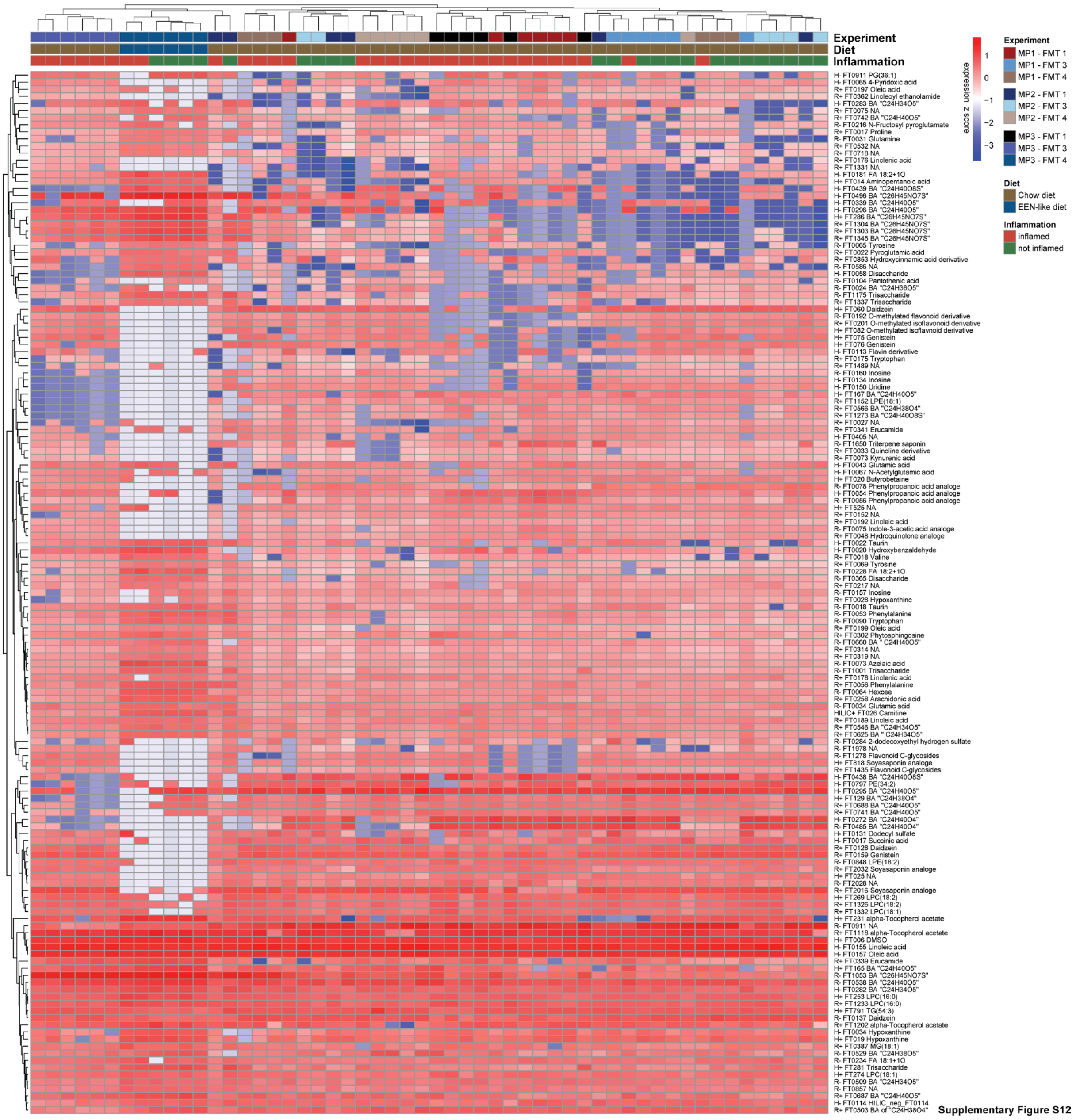


Supplementary Figure S12

***Figure S12: Untargeted metabolomics in all transfer experiments***

*Differential heat map (based on Wilcoxon test) of untargeted metabolomics measured in Il10$^{-/-}$ mice from MP1, 2 (excluding relapse samples) and 3. Samples clustered according to inflammation and their similarity of metabolite features. Color codes on top indicate from which experiment the mice belong to, what diet they had (chow, EEN-like), their classification of inflammation (histopathology score > 3 was considered as inflamed phenotype)*

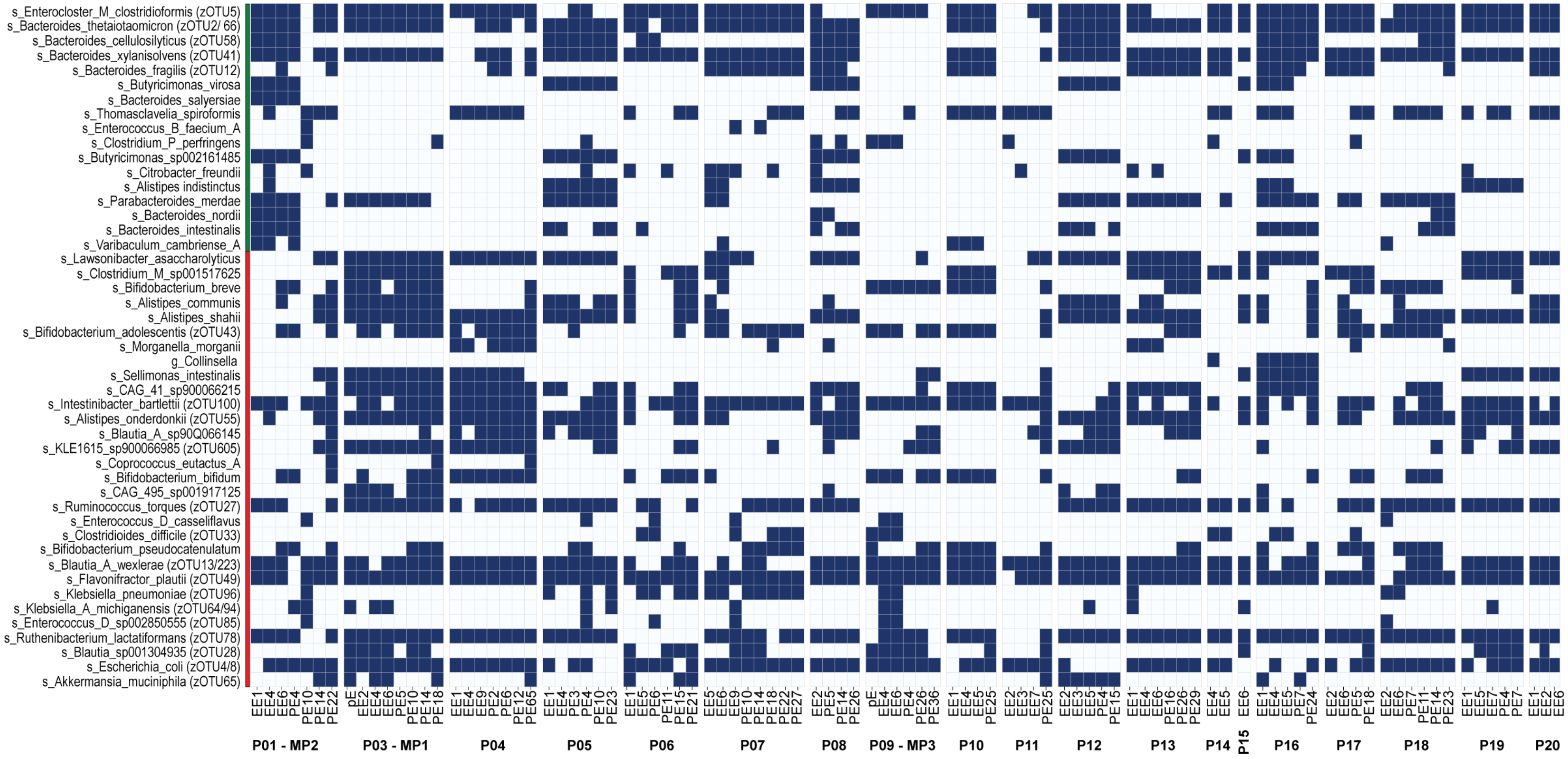

***Figure S13: Abundance of differentially abundant strains from transfer experiments in CD-EEN patients***

*Abundance of differentially abundant strains from* ***Figure 5E*** *in all CD-EEN patients (blue = abundant, grey = not detected). Strain abundance cut-off 20%, species abundance cut-off* $10^{-5}$*, prevalence ≥ 1.*

**A**

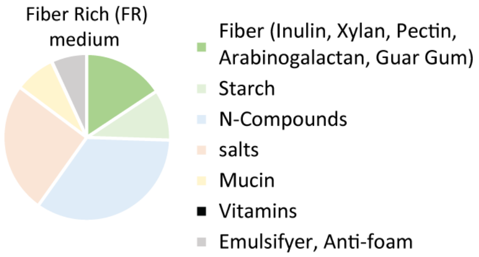


**B**

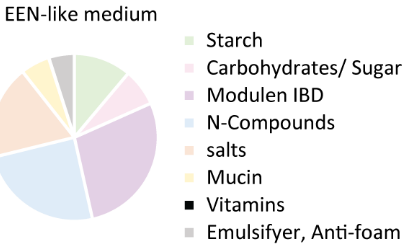


**C**

| | Component | g/L FR (MacFarlane et al. 1998) | g/L EEN-like |
|---|---|---|---|
| Fiber | Pectin (from citrus) | 2 | |
| | Guar gum | 1 | |
| | Xylan (from oatspelt) | 2 | |
| | Inulin (from *Dahlia* tubers) | 1 | |
| | Arabinogalactan (larch wood) | 2 | |
| Starch | Starch (wheat) | 5 | |
| | Starch (rice) | | 7.9 |
| Carbohydrates/ Sugar | Sucrose | | 1.3 |
| | Glucose | | 3.6 |
| | Fructose | | 0.2 |
| Mudulen IBD ® | Modulen IBD | | 20 |
| N-Compunds | Tryptone | 5 | 5 |
| | Peptone | 5 | 5 |
| | Yeast Extract | 4.5 | 4.5 |
| | Casein | 3 | 3 |
| Salts | NaCl | 4.5 | 4.5 |
| | KCl | 4.5 | 4.5 |
| | $MgSO_4 \cdot 7H_20$ | 1.25 | 1.25 |
| | $CaCl_2$ | 0.113 | 0.113 |
| | $NaHCO_3$ | 1.5 | 1.5 |
| | $KH_2PO_4$ | 0.5 | 0.5 |
| | L-Cysteine HCl | 0.8 | 0.8 |
| | $FeSO_4 \cdot 7H_20$ | 0.005 | 0.005 |
| | Bile salts (no 3) | 0.4 | 0.4 |
| Mucin | Porcine gastric mucin (type II) | 4 | 4 |
| Vitamin solution (Gibson and Wang 1994) | Pantothenate | 10 mg/L | 10 mg/L |
| | Nicotinamide | 5 mg/L | 5 mg/L |
| | Thiamine | 4 mg/L | 4 mg/L |
| | Biotin | 2 mg/L | 2 mg/L |
| | B12 | 0.5 mg/L | 0.5 mg/L |
| | Menadione | 1 mg/L | 1 mg/L |
| | PABA | 5 mg/L | 5 mg/L |
| | Hemin | 10 mg/L | 10 mg/L |
| Emulsifyer, Anti-foam | Anti-foam | 2.5 mL | 2.5 mL |
| | Tween 80 | 1 mL | 1 mL |

**D**

| | Chow | EEN-like |
|---|---|---|
| **Fiber [g/kg]** | 170 | 50 |
| **Fiber Source** | soy plant | cellulose |
| **Protein [E%]** | 27 | 23 |
| **Carbohydrataes [E%]** | 61 | 64 |
| **Sucrose [g/kg]** | 0 | 50 |
| **Fat [E%]** | 12 | 13 |
| **kcal/kg** | 3338 | 3660 |

***Figure S14: Composition of used diets and media in model systems***

***A, B:*** *Pie chart of gut chemostat medium composition.* ***A*** *FR medium.* ***B*** *EEN-like medium.* ***C*** *Composition of gut chemostat medium (g/L) of FR and EEN-like medium.* ***D*** *Composition of mouse diets (chow and EEN-like mouse diet) in Energy % (E%) or g/kg. EEN-like diet = SNIFF S5745-E90.*
